# Prevalence and characteristics of genetic disease in adult kidney stone formers

**DOI:** 10.1101/2023.07.23.23292924

**Authors:** Manuel A. Anderegg, Eric G. Olinger, Matteo Bargagli, Rob Geraghty, Lea Pohlmeier, Alexander Nater, Rémy Bruggmann, John A. Sayer, Bruno Vogt, André Schaller, Daniel G. Fuster

## Abstract

**Background:** Molecular mechanisms of kidney stone formation remain unknown in most patients. Previous studies showed high heritability of nephrolithiasis, but data on prevalence and characteristics of genetic disease in unselected adults with nephrolithiasis are lacking.

**Methods:** We performed whole exome sequencing in 787 participants of the Bern Kidney Stone Registry, an unselected cohort of adults with ≥ 1 past kidney stone episode (KSF), and 114 non-stone-forming individuals (NKSF). A total of 34 established nephrolithiasis genes were analyzed and variants assessed according to ACMG criteria. Pathogenic (P) or likely pathogenic (LP) variants were considered diagnostic.

**Results:** Mean age of KSF was 47±15 years, and 18 % were first time KSF. A Mendelian kidney stone disease was present in 2.9% (23 of 787) of KSF. The most common genetic diagnoses were cystinuria (*SLC3A1*, *SLC7A9*; n=13), Vitamin D-24 hydroxylase deficiency (*CYP24A1*; n=5) and primary hyperoxaluria (*AGXT, GRHPR, HOGA1*; n=3). 8.1% (64 of 787) of KSF were monoallelic for LP/P variants predisposing to nephrolithiasis, most frequently in *SLC34A1/A3* or *SLC9A3R1* (n=37), *CLDN16* (n=8) and *CYP24A1* (n=8). KSF with Mendelian disease had a lower age at the first stone event (30±14 years vs. 36±14 years, p=0.003), were more likely to have cystine stones (23.4 % vs. 1.4 %) and less likely to have calcium oxalate monohydrates stones (31.9 % vs. 52.5 %) compared to KSF without genetic diagnosis. The phenotype of KSF with variants predisposing to nephrolithiasis was subtle and showed significant overlap with KSF without diagnostic variants. In NKSF, no Mendelian disease was detected, and LP/P variants were significantly less prevalent compared to KSF (1.8 % vs. 8.1%).

**Conclusion:** Mendelian disease is uncommon in unselected adult KSF, yet variants predisposing to nephrolithiasis are significantly enriched in adult KSF.

## INTRODUCTION

Nephrolithiasis is a common global healthcare problem ^1^. Kidney stones recur frequently and cause substantial morbidity, reduced quality of life and enormous cost ^2-4^. Most kidney stones are classified as “idiopathic”, indicating that the pathogenesis is unclear. Consequently, undifferentiated dietary and pharmacological preventive measures are initiated, but many patients continue to form stones ^5,6^. Hence, our current strategies in managing this very common and relapsing disease are clearly inadequate. There is an unmet need for novel diagnostic and therapeutic approaches.

Kidney stone formation is strongly influenced by genetic factors: a positive family history is present in 30-60% of individuals with kidney stones ^7^, and both twin ^8^ and genealogy ^9^ studies revealed a high heritability of nephrolithiasis. More than 30 monogenic, Mendelian causes of nephrolithiasis have been described thus far ^10^. Recent studies have also highlighted the importance of intermediate effect size / incomplete penetrance variants in increasing the risk for nephrolithiasis^11-14^. Identification of patients with Mendelian forms of nephrolithiasis remains a major challenge, and no clear consensus exists on clinical parameters to guide genetic testing, although extreme clinical phenotypes usually prompt genetic testing. As a result, many cases of Mendelian forms of nephrolithiasis are missed and incorrectly labelled “idiopathic”. Whole exome sequencing (WES) is widely applied as diagnostic tool for rare diseases and the detection of pathogenic variants in cancer ^15,16^. In contrast, the diagnostic utility of WES has not been established for most constitutional disorders, such as nephrolithiasis. In small, selected cohorts of early-onset or familial nephrolithiasis, a Mendelian cause was identified in 6.8 - 29.4% of cases ^10,17-19^. However, the frequency and phenotypic spectrum of genetic disease in sporadic adult-onset nephrolithiasis, by far the most common type encountered in clinical routine, is unknown. To address these important knowledge gaps, we employed WES in a deeply phenotyped, unselected European cohort of 787 adult kidney stone formers (KSF) and in 114 non-stone forming individuals (NKSF).

## METHODS

### Study cohort

The study was conducted with participants enrolled in the Bern Kidney Stone Registry (BKSR), an. observational cohort of adult kidney stone formers described previously and in the supplemental information^20-23^. Inclusion criteria for the BKSR are i) written informed consent, ii) age ≥ 18 years, and iii) ≥ 1 past kidney stone episode.

Inclusion criteria for NKSF were i) written informed consent, ii) age ≥ 18 years, and iii) no history of past kidney stones and no evidence of asymptomatic nephrolithiasis or nephrocalcinosis on ultrasound at enrolment.

### Whole exome sequencing and variant calling

Isolation of genomic DNA, exome capture, high-throughput sequencing and bioinformatic analysis including joint variant calling using Genome analysis Toolkit v3.8 (GATK) according to GATK best practices recommendations^24,25^ and annotation using Ensembl Variant Effect Predictor Release 106 (VEP) ^26^ was performed using established methods, described in more detail in Supplemental Methods. After filtering for predicted consequences and genome aggregation database (gnomAD) minor allele frequency (<1% in all populations) ^27^, variants in 34 genes ^10,17-19,28^ previously implicated in Mendelian kidney stone disease were examined (Fig. 1, lower panel). Further variant stratification was performed according to the recommendations of the American College of Medical Genetics and Genomics (ACMG)^29^ after manual review of phenotype data. Only variants classified as likely pathogenic (LP) or pathogenic (P) applying the ACMG criteria were defined as diagnostic variants leading to a genetic diagnosis, as Mendelian or as LP/P variants predisposing to nephrolithiasis (LP/P variants) depending on evidence available from this study and/or from published case/control studies.^11,12,30,31^ (Fig. 1 and Supplemental Methods).

**Figure 1:**
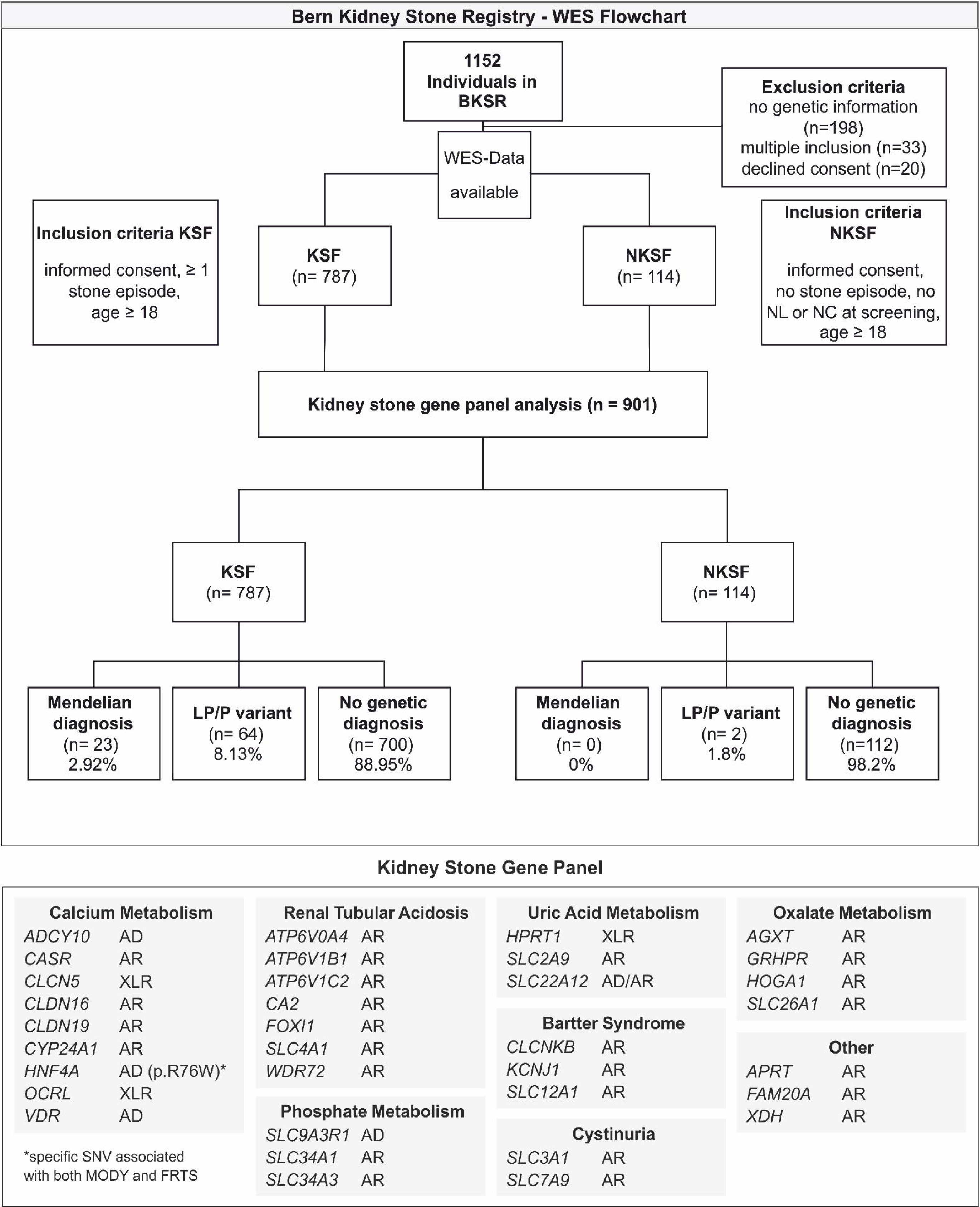
Upper Panel: Flowchart of patient inclusion, exclusion and genetic analysis in the Bern Kidney Stone Registry (BKSR). Data of 1152 individuals recruited into the BKSR were analyzed. After exclusion of individuals without genetic information available, declined consent or multiple inclusions, the analyzed cohort consisted of 901 individuals (787 stone formers (KSF) and 114 non stone forming controls (NKSF). Before inclusion in the BKSR, NKSF underwent ultrasound imaging to exclude nephrolithiasis (NL) and/or nephrocalcinosis (NC). Genetic analysis was performed in a standardized prioritization pathway in 34 known kidney stone genes. N: number of individuals. Genetic Diagnosis: Likely pathogenic or pathogenic variant according to ACMG criteria. LP/P Variant: monoallelic LP/P variant, predisposing to nephrolithiasis. WES: whole exome sequencing. **Lower Panel: Kidney stone disease gene panel used for genetic analysis.** Panel of 34 known nephrolithiasis genes used with their inheritance mode, as accepted for classification as “Mendelian disease” in this manuscript, grouped by phenotypes. AD: autosomal dominant, AR: autosomal recessive, XLR: X-linked recessive, MODY: maturity-onset diabetes of the young, FRTS: Fanconi renal tubular syndrome.

### Statistical Analysis

Continuous variables were reported as medians with 25^th^-75^th^ percentiles or means with standard deviations and categorical variables were reported as counts with percentages, as appropriate. Different statistical methods were employed to analyze differences between independent groups, depending on the nature of the variables involved. Mann-Whitney U test was used for non-normally distributed continuous variables, while Student’s t-test was applied for normally distributed variables. For categorical variables, the Fisher’s Exact Test was implemented. Findings from these analyses were summarized and presented as descriptive tables. Statistical tests were two-sided and a *p*-value < 0.05 was considered statistically significant. Statistical analyses were performed using Stata, version 16 (Statacorp, TX, USA). Pie-charts and dot plots were generated with GraphPad Prism, version 8.4.3 (GraphPad Software, San Diego, CA, USA).

## RESULTS

### Study cohort

We performed WES in a total of 901 individuals, including 787 KSF and 114 NKSF (Fig. 1). Clinical characteristics of the study cohort, separated in KSF and NKSF, are shown in Table 1. Mean age ± standard deviation (SD) was 46.6±14.5 years in KSF and 42.4±14.9 years in NKSF, respectively. The majority of KSF were male (71.8 %, n=565), whereas the percentage of men was lower in NKSF (54 %, n=61). KSF were significantly more likely to report a positive family history of kidney stones compared to NKSF (43.6 % vs. 6.8 %, Table 1).

**Table 1.** Characteristics of the study cohort. Characteristics are indicated separately for participants with and without kidney stone disease. Categorical variables are described by number of participants N (%), continuous variables are described by their mean (SD) or median (25th-75th percentile). eGFR, estimated glomerular filtration rate; BSA, body surface area; PTH, parathyroid hormone; SD, standard deviation. * The Non kidney stone former (NKSF) with a diagnostic variant (#5948) has been removed for phenotypic analyses.

| Characteristics | Kidney stone formers<br>(N=787) | Non kidney stone<br>formers (N=113) * | p-values |
| --- | --- | --- | --- |
| Age, years | 46.64 (14.57) | 42.41 (14.88) | 0.007 |
| Males | 565 (71.8%) | 61 (54.0%) | <0.001 |
| Body mass index, kg/m<sup>2</sup> | 26.83 (5.08) | 26.34 (4.55) | 0.37 |
| Hypertension | 255 (36.0%) | 28 (35.9%) | 1.00 |
| Diabetes | 24 (3.1%) | 1 (1.1%) | 0.50 |
| Obesity, BMI > 30 kg/m<sup>2</sup> | 168 (22.4%) | 14 (15.1%) | 0.11 |
| Hyperuricemia | 346 (44.0%) | 54 (47.8%) | 0.48 |
| Family history of kidney stone disease | 317 (43.6%) | 3 (6.8%) | <0.001 |
| Kidney stone recurrence, > 1 stone event | 614 (82.2%) |  |  |
| Age at first stone event, years | 35.41 (13.85) |  |  |
| Total number of stone events | 3.00 (2.00, 4.00) |  |  |
| **Blood parameters** |  |  |  |
| Total calcium, mmol/L | 2.36 (0.12) | 2.30 (0.12) | <0.001 |
| Ionized calcium, mmol/L | 1.21 (1.19, 1.23) | 1.19 (1.18, 1.22) | 0.002 |
| Phosphorus, mmol/L | 1.01 (0.17) | 1.01 (0.16) | 0.89 |
| Magnesium, mmol/L | 0.83 (0.07) | 0.86 (0.08) | <0.001 |
| Uric acid, umol/L | 328.96 (169.56) | 308.44 (71.51) | 0.26 |
| Intact PTH, ng/L | 39.06 (30.00, 51.00) | 37.50 (30.50, 48.00) | 0.56 |
| 25-OH Vitamin D<sub>3</sub>, nmol/L | 40.18 (27.00, 59.00) | 48.50 (32.72, 72.44) | 0.017 |
| eGFR creatinine Equation CKD-EPI 2009, mL/min per 1.73 m<sup>2</sup> BSA | 94.18 (21.08) | 96.31 (21.88) | 0.36 |
| **Urine parameters** |  |  |  |
| Total urine volume, L/24h | 1.98 (1.47, 2.54) | 2.10 (1.49, 2.65) | 0.41 |
| Urine pH | 5.94 (5.39, 6.60) | 6.00 (5.50, 6.75) | 0.14 |
| Urine sodium / creatinine ratio, mmol/mmol/24h | 13.62 (10.97, 16.98) | 13.76 (9.92, 16.21) | 0.37 |
| Urine potassium / creatinine ratio, mmol/mmol/24h | 4.70 (3.77, 5.82) | 5.56 (4.43, 6.55) | <0.001 |
| Urine uric acid / creatinine ratio, mmol/mmol/24h | 0.61 (0.10) | 0.38 (0.27) | <0.001 |
| Urine calcium / creatinine ratio, mmol/mmol/24h | 0.43 (0.30, 0.58) | 0.33 (0.21, 0.44) | <0.001 |
| Urine magnesium / creatinine ratio, mmol/mmol/24h | 0.30 (0.24, 0.38) | 0.32 (0.27, 0.42) | 0.059 |
| Urine citrate / creatinine ratio, mmol/mmol/24h | 0.19 (0.13, 0.27) | 0.24 (0.18, 0.30) | 0.003 |
| Urine phosphate / creatinine ratio, mmol/mmol/24h | 2.18 (0.54) | 2.24 (0.57) | 0.41 |
| Urine oxalate / creatinine ratio, mmol/mmol/24h | 0.03 (0.02, 0.04) | 0.03 (0.02, 0.05) | 0.062 |
| **Stone phenotypes** |  |  |  |
| Calcium oxalate dihydrate | 73 (12.9%) |  |  |
| Calcium oxalate monohydrate | 287 (50.8%) |  |  |
| Calcium phosphate | 130 (23.0%) |  |  |
| Uric acid | 38 (6.7%) |  |  |
| Cystine | 18 (3.2%) |  |  |
| Struvite | 15 (2.7%) |  |  |

### Genetic analysis

Among 787 unrelated, unselected adult KSF, we discovered in 23 patients (2.9 %) a Mendelian form of nephrolithiasis, i.e., pathogenic (P) or likely pathogenic (LP) variants in genes associated with Mendelian forms of nephrolithiasis respecting the relevant mode of inheritance (autosomal dominant and/or autosomal recessive and X-linked recessive) (Table 2, Fig. 1, Fig. 2). Mendelian disease was detected in nine of 34 analyzed genes (n= number of patients): *AGXT* (n=1)*, ATP6V1B1* (n=1)*, CYP24A1* (n=5)*, GRHPR* (n=1)*, HOGA1* (n=1)*, SLC3A1* (n=9)*, SLC4A1* (n=1)*, SLC7A9* (n=4)*, SLC12A1* (n=1).

**Figure 2:**
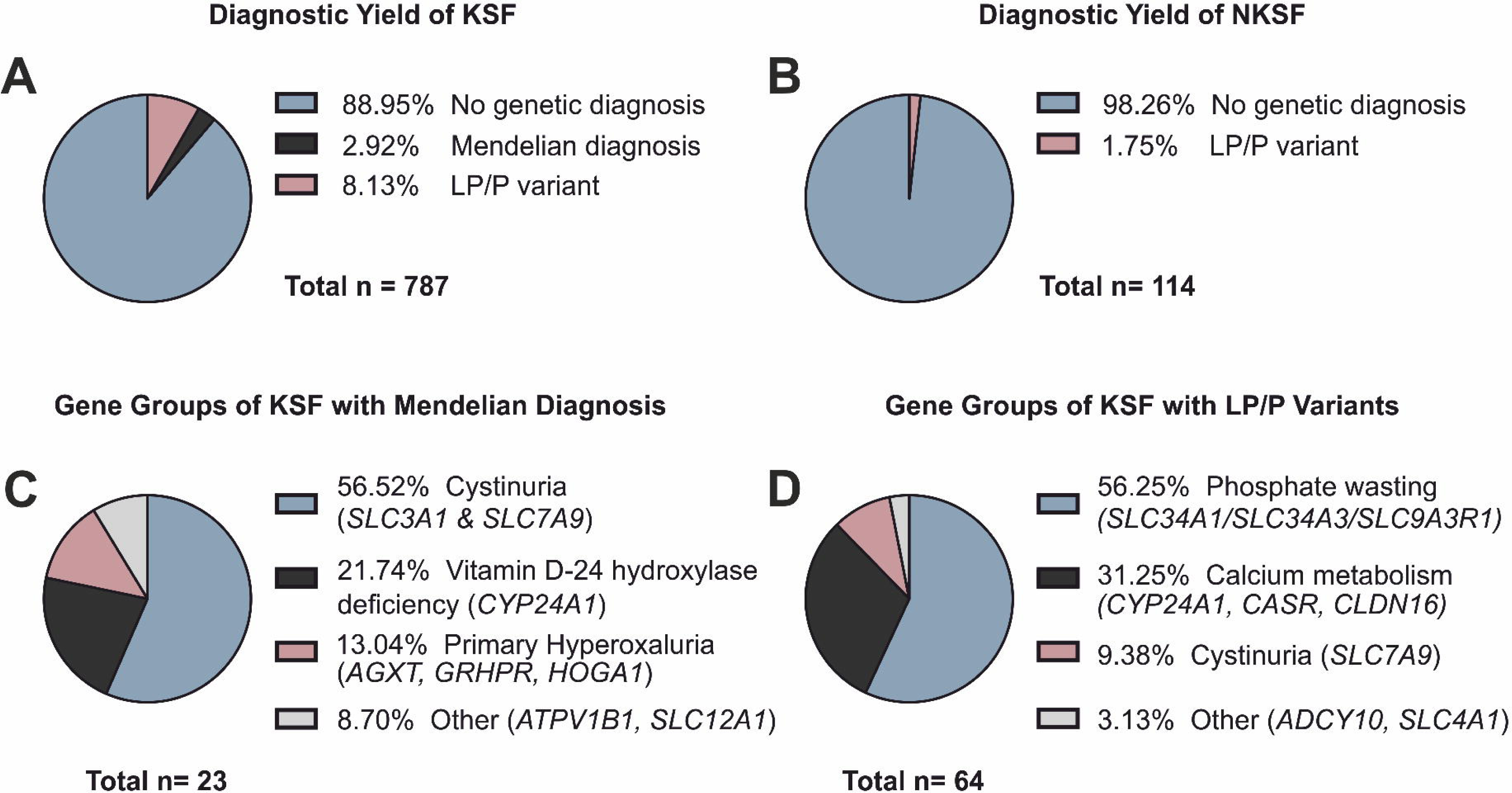
Overall yield of genetic diagnoses. (A) Diagnostic yield (Mendelian vs. LP/P variants predisposing to nephrolithiasis)) in kidney stone formers (KSF), (B) Diagnostic yield in non-stone forming controls (NKSF), C) Overview of Mendelian diagnoses in KSF, sorted by phenotype groups, (D) Overview of LP/P variants predisposing to nephrolithiasis in KSF, sorted by phenotype groups.

**Table 2:**
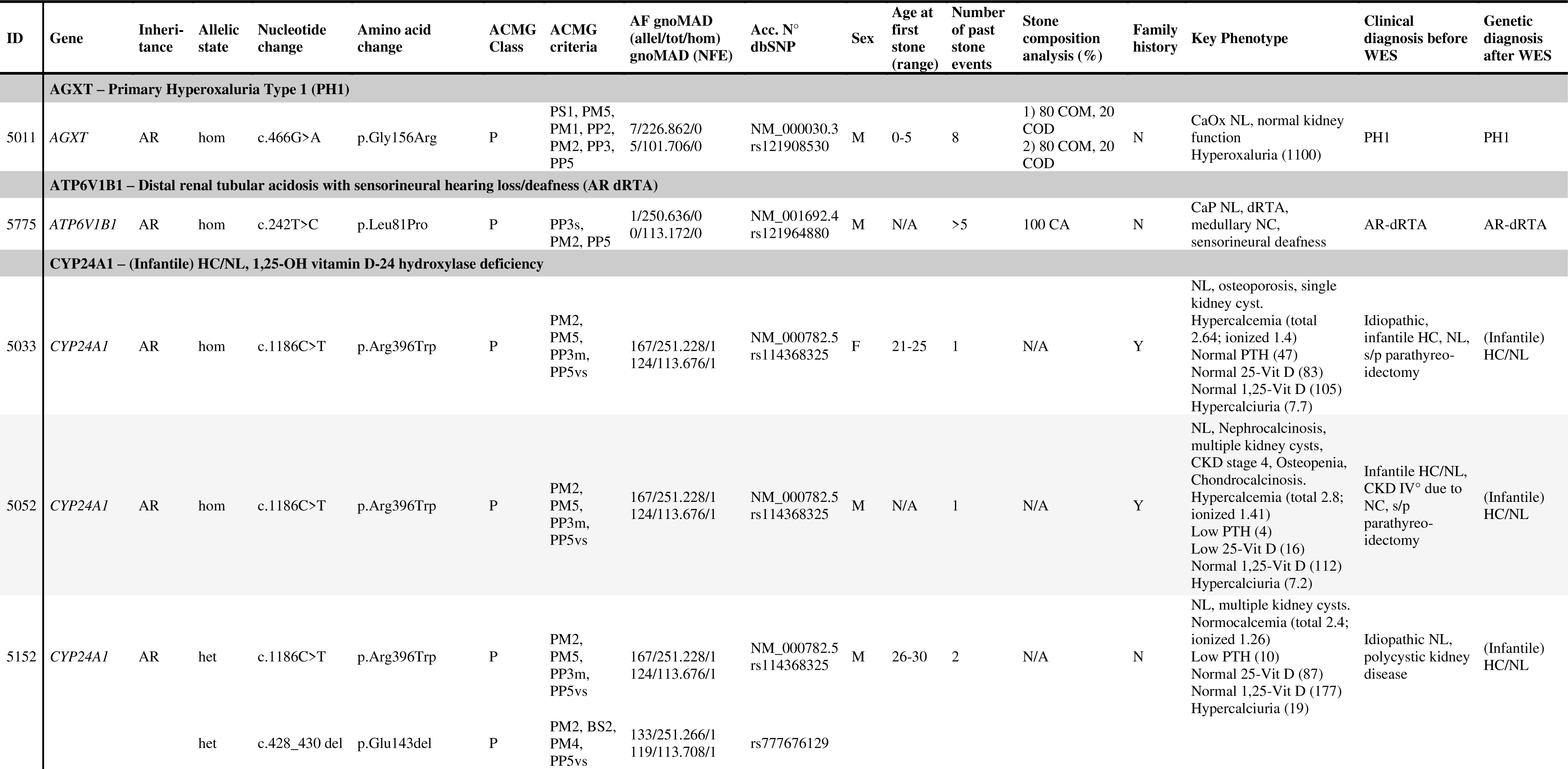
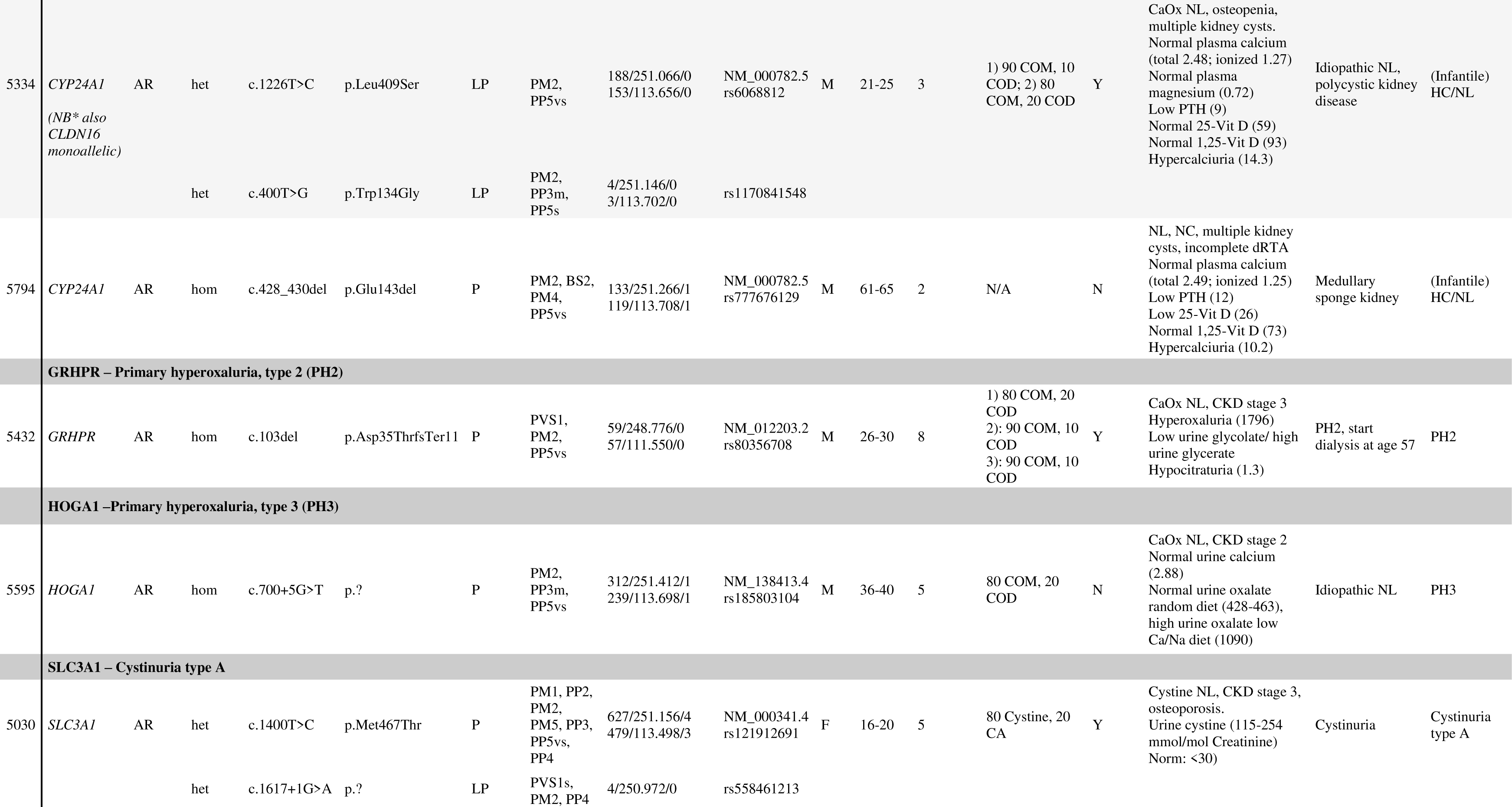
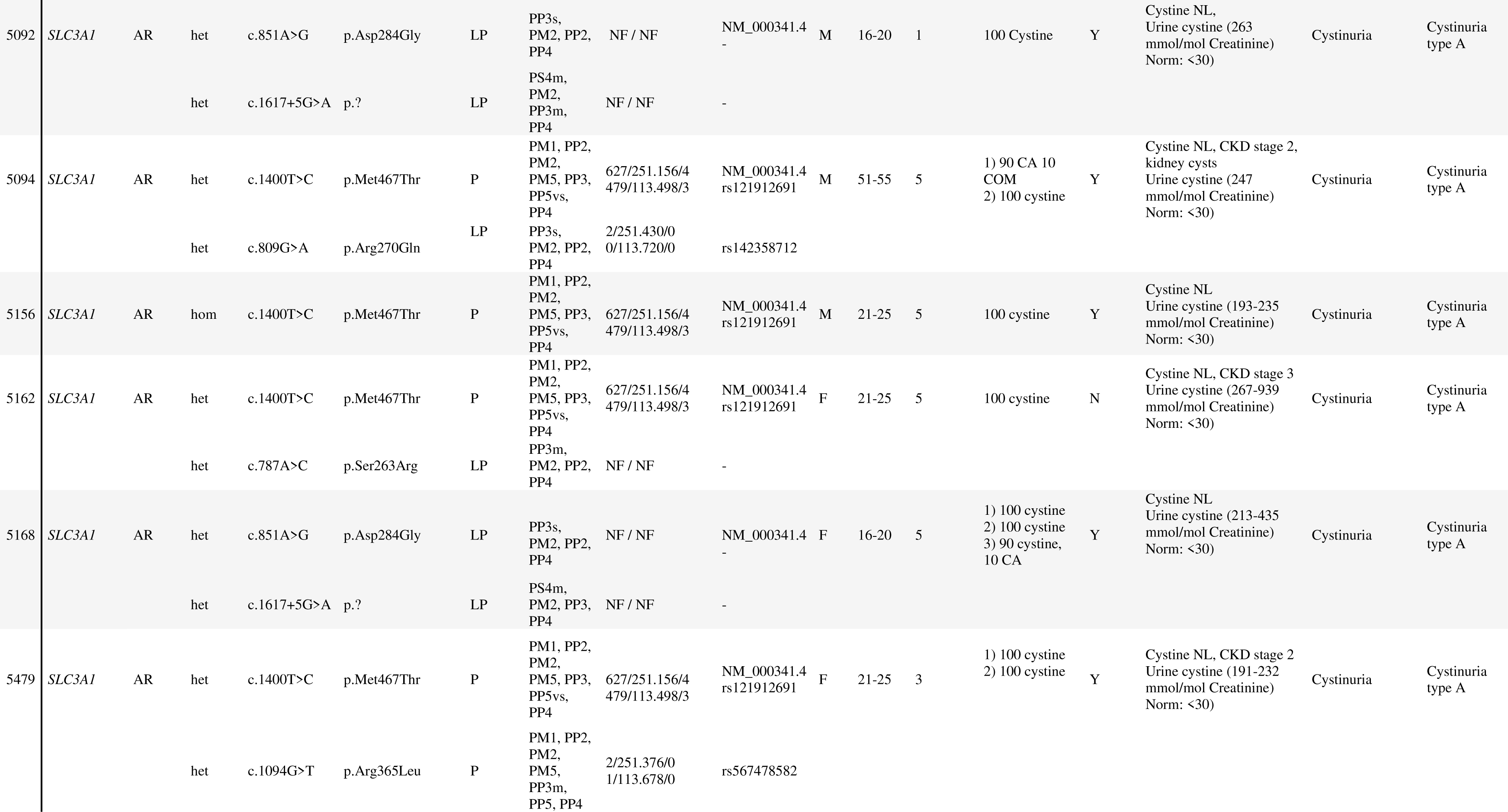
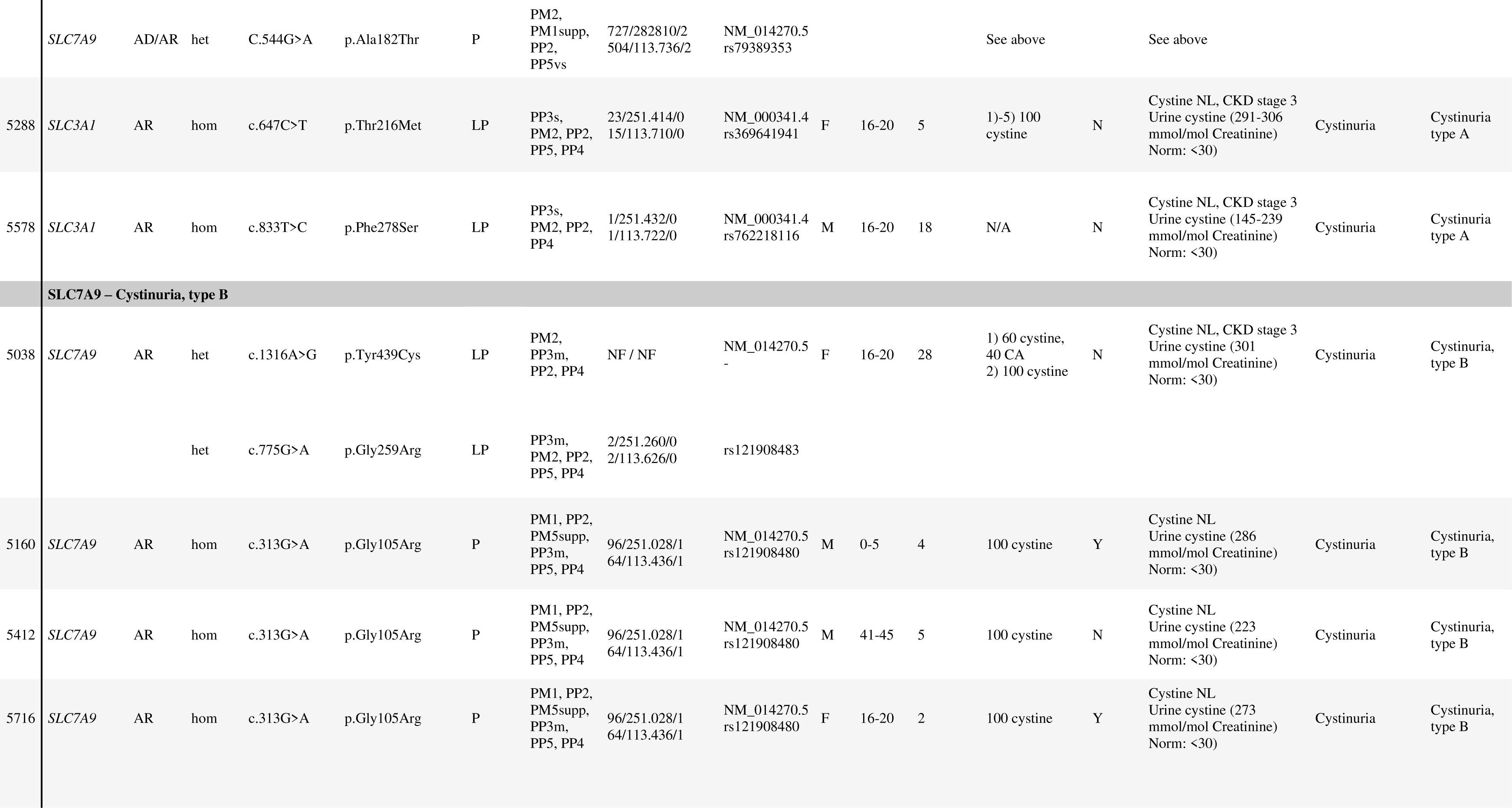
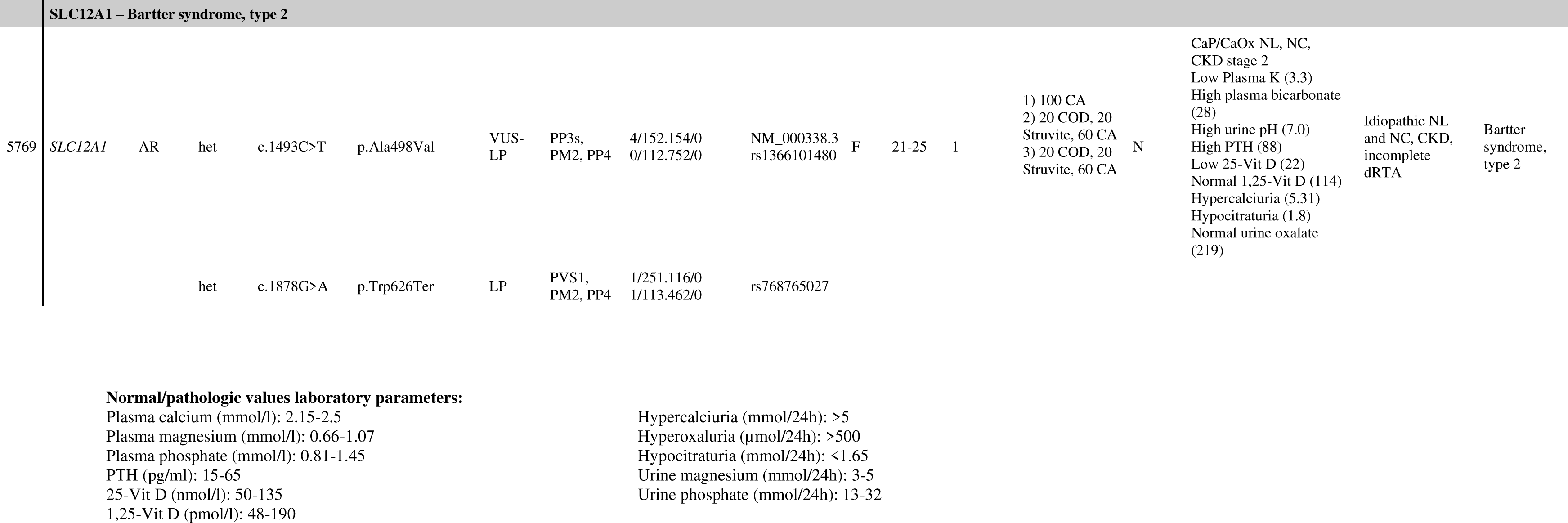
Genotype and phenotype of solved patients with Mendelian (monogenic) diagnosis. Acc.N°: RefSeq accession number, ACMG: American College of Medical Genetics, AF: allele frequency, AR: autosomal recessive, AD: autosomal dominant, dbSNP: Reference SNP number of variant, het: heterozygous, hom: homozygous, LP: likely pathogenic, P: pathogenic, tot: total, Novel: mutation detected for the first time in this study/not previously described, NS: nonsense, Family history: N= no, Y=yes, M: male, F: female, mo: months, NL: Nephrolithiasis, NC: Nephrocalcinosis, dRTA: distal renal tubular acidosis, PHPTH: primary hyperparathyroidism, CKD: chronic kidney disease. Stone analysis: CaOx: Calcium oxalate, COM: Calcium oxalate monohydrate, COD: Calcium oxalate dihydrate, CaP: Calcium phosphate, CA: Carbonate apatite, Brushite: Calcium hydrogen phosphate dihydrate, MAP: Magnesium ammonium phosphate (=Struvite), UA: Uric acid.

A total of 66 individuals (8.4%), 64 (8.1%) of which were KSF, presented with LP/P variants not fulfilling our stringent criteria for Mendelian disease, but predisposing for nephrolithiasis (Supplementary Table 1). LP/P variants were detected in nine of 34 genes (n=number of patients): *ADCY10* (n=1), *CASR* (n=4), *CLDN16* (n=8*), CYP24A1* (n=8)*, SLC4A1* (n=1)*, SLC7A9* (n=3)*, SLC34A1* (n=15)*, SLC34A3* (n=17) and *SLC9A3R1* (n=8). Of these variants, 18 % (12 of 66) were novel, previously unreported in ClinVar or HGMD (Supplementary Table 2).

Prior to WES, a Mendelian form of nephrolithiasis was known or suspected in 18 of 23 individuals (78 %) with a post WES Mendelian diagnosis, despite each case being reviewed by a kidney stone expert. Therefore, in five of 23 individuals (22 %) genetic analysis established a new or corrected a suspected *a priori* diagnosis. The diagnostic yield was similar between first-time and recurrent stone formers (Supplementary Table 3), and between men and women (Table 3, Supplementary Table 4).

**Table 3.**
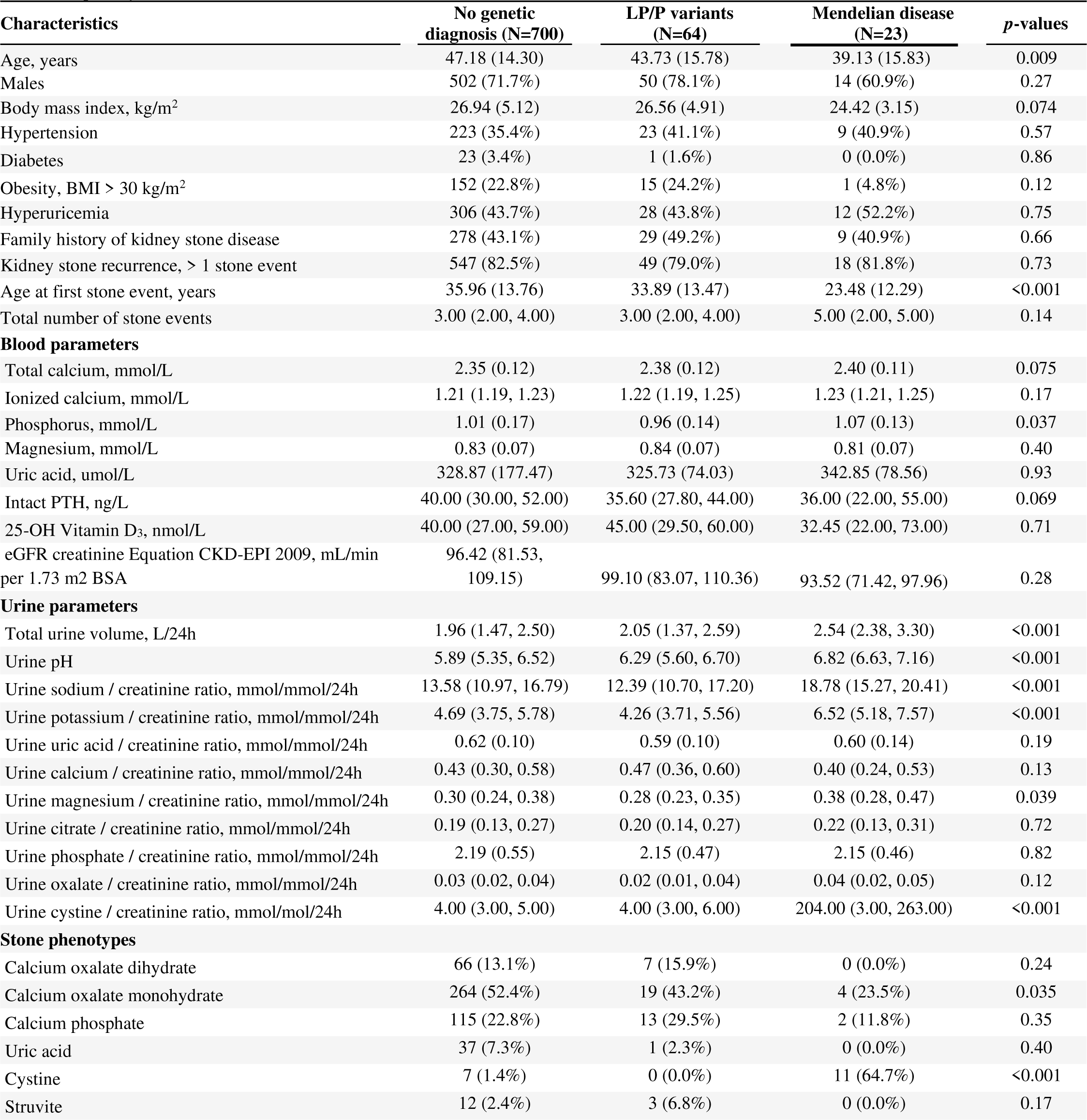
Characteristics of stone formers with and without pathogenic gene variants. Characteristics are indicated separately for stone formers with Mendelian disease and with/without LP/P predisposing gene variants. Categorical variables are described by number of participants N (%), continuous variables are described by their mean (SD) or median (25th-75th percentile). eGFR, estimated glomerular filtration rate; BSA, body surface area; PTH, parathyroid hormone; SD, standard deviation.

| Characteristics | No genetic diagnosis (N=700) | LP/P variants (N=64) | Mendelian disease (N=23) | *p*-values |
| --- | --- | --- | --- | --- |
| Age, years | 47.18 (14.30) | 43.73 (15.78) | 39.13 (15.83) | 0.009 |
| Males | 502 (71.7%) | 50 (78.1%) | 14 (60.9%) | 0.27 |
| Body mass index, kg/m<sup>2</sup> | 26.94 (5.12) | 26.56 (4.91) | 24.42 (3.15) | 0.074 |
| Hypertension | 223 (35.4%) | 23 (41.1%) | 9 (40.9%) | 0.57 |
| Diabetes | 23 (3.4%) | 1 (1.6%) | 0 (0.0%) | 0.86 |
| Obesity, BMI > 30 kg/m<sup>2</sup> | 152 (22.8%) | 15 (24.2%) | 1 (4.8%) | 0.12 |
| Hyperuricemia | 306 (43.7%) | 28 (43.8%) | 12 (52.2%) | 0.75 |
| Family history of kidney stone disease | 278 (43.1%) | 29 (49.2%) | 9 (40.9%) | 0.66 |
| Kidney stone recurrence, > 1 stone event | 547 (82.5%) | 49 (79.0%) | 18 (81.8%) | 0.73 |
| Age at first stone event, years | 35.96 (13.76) | 33.89 (13.47) | 23.48 (12.29) | <0.001 |
| Total number of stone events | 3.00 (2.00, 4.00) | 3.00 (2.00, 4.00) | 5.00 (2.00, 5.00) | 0.14 |
| **Blood parameters** |  |  |  |  |
| Total calcium, mmol/L | 2.35 (0.12) | 2.38 (0.12) | 2.40 (0.11) | 0.075 |
| Ionized calcium, mmol/L | 1.21 (1.19, 1.23) | 1.22 (1.19, 1.25) | 1.23 (1.21, 1.25) | 0.17 |
| Phosphorus, mmol/L | 1.01 (0.17) | 0.96 (0.14) | 1.07 (0.13) | 0.037 |
| Magnesium, mmol/L | 0.83 (0.07) | 0.84 (0.07) | 0.81 (0.07) | 0.40 |
| Uric acid, umol/L | 328.87 (177.47) | 325.73 (74.03) | 342.85 (78.56) | 0.93 |
| Intact PTH, ng/L | 40.00 (30.00, 52.00) | 35.60 (27.80, 44.00) | 36.00 (22.00, 55.00) | 0.069 |
| 25-OH Vitamin D<sub>3</sub>, nmol/L | 40.00 (27.00, 59.00) | 45.00 (29.50, 60.00) | 32.45 (22.00, 73.00) | 0.71 |
| eGFR creatinine Equation CKD-EPI 2009, mL/min per 1.73 m<sup>2</sup> BSA | 96.42 (81.53, 109.15) | 99.10 (83.07, 110.36) | 93.52 (71.42, 97.96) | 0.28 |
| **Urine parameters** |  |  |  |  |
| Total urine volume, L/24h | 1.96 (1.47, 2.50) | 2.05 (1.37, 2.59) | 2.54 (2.38, 3.30) | <0.001 |
| Urine pH | 5.89 (5.35, 6.52) | 6.29 (5.60, 6.70) | 6.82 (6.63, 7.16) | <0.001 |
| Urine sodium / creatinine ratio, mmol/mmol/24h | 13.58 (10.97, 16.79) | 12.39 (10.70, 17.20) | 18.78 (15.27, 20.41) | <0.001 |
| Urine potassium / creatinine ratio, mmol/mmol/24h | 4.69 (3.75, 5.78) | 4.26 (3.71, 5.56) | 6.52 (5.18, 7.57) | <0.001 |
| Urine uric acid / creatinine ratio, mmol/mmol/24h | 0.62 (0.10) | 0.59 (0.10) | 0.60 (0.14) | 0.19 |
| Urine calcium / creatinine ratio, mmol/mmol/24h | 0.43 (0.30, 0.58) | 0.47 (0.36, 0.60) | 0.40 (0.24, 0.53) | 0.13 |
| Urine magnesium / creatinine ratio, mmol/mmol/24h | 0.30 (0.24, 0.38) | 0.28 (0.23, 0.35) | 0.38 (0.28, 0.47) | 0.039 |
| Urine citrate / creatinine ratio, mmol/mmol/24h | 0.19 (0.13, 0.27) | 0.20 (0.14, 0.27) | 0.22 (0.13, 0.31) | 0.72 |
| Urine phosphate / creatinine ratio, mmol/mmol/24h | 2.19 (0.55) | 2.15 (0.47) | 2.15 (0.46) | 0.82 |
| Urine oxalate / creatinine ratio, mmol/mmol/24h | 0.03 (0.02, 0.04) | 0.02 (0.01, 0.04) | 0.04 (0.02, 0.05) | 0.12 |
| Urine cystine / creatinine ratio, mmol/mol/24h | 4.00 (3.00, 5.00) | 4.00 (3.00, 6.00) | 204.00 (3.00, 263.00) | <0.001 |
| **Stone phenotypes** |  |  |  |  |
| Calcium oxalate dihydrate | 66 (13.1%) | 7 (15.9%) | 0 (0.0%) | 0.24 |
| Calcium oxalate monohydrate | 264 (52.4%) | 19 (43.2%) | 4 (23.5%) | 0.035 |
| Calcium phosphate | 115 (22.8%) | 13 (29.5%) | 2 (11.8%) | 0.35 |
| Uric acid | 37 (7.3%) | 1 (2.3%) | 0 (0.0%) | 0.40 |
| Cystine | 7 (1.4%) | 0 (0.0%) | 11 (64.7%) | <0.001 |
| Struvite | 12 (2.4%) | 3 (6.8%) | 0 (0.0%) | 0.17 |

### Genotype-phenotype correlation

Mean age of first kidney stone event (23.5±12.3 years vs. 35.9±13.8 years, p<0.001) and mean age of presentation for metabolic-work up (39±15.8 years vs. 47.2±14.3 years, p=0.009) were lower in KSF with Mendelian diagnosis compared to KSF without, with intermediate results for patients with LP/P variants predisposing for nephrolithiasis (Table 3, Supplemental Fig. 3). Furthermore, cystine stones were significantly more common (64.7 % vs. 1.4 %) and calcium oxalate monohydrates stones less common (23.5 % vs. 52.5 %) in KSF with Mendelian diagnosis compared to KSF with LP/P variants or without genetic diagnosis, in line with higher urinary cystine and urine pH in KSF with Mendelian diagnosis (Table 3, Supplemental Fig. 2). The prevalence of a positive family history of kidney stone disease was similar in all three groups (43.1 % vs. 49.2% vs. 46.0 %). Additionally, KSF with Mendelian disease had more past kidney stone events [5 (2,5) vs. 3 (2,4), p=0.052] than KSF with LP/P variants. In contrast, KSF with LP/P variants had significantly lower plasma phosphate (Table 3).

### Mendelian disease

The most common Mendelian diagnosis was cystinuria (n=13) due to biallelic diagnostic variants in *SLC3A1* (type A cystinuria, n=9) or *SLC7A9* (type B cystinuria, n=4). Urine cystine excretion, where available, was increased in all KSF with genetic diagnosis of cystinuria. One patient had biallelic diagnostic variants in *SLC3A1* and a monoallelic LP/P variant in *SLC7A9*. KSF with biallelic diagnostic *SLC3A1* or *SLC7A9* variants all had cystine stones.

Five patients had biallelic diagnostic variants in *CYP24A1,* which encodes the Vitamin-D inactivating enzyme Vitamin D-24 hydroxylase. Interestingly, all KSF with biallelic variants in *CYP24A1* (but no KSF with a monoallelic *CYP24A1* variant) displayed cystic kidney disease, as reported previously ^31,32^. Common misdiagnoses for patients with biallelic variants in *CYP24A1* (n=3) were polycystic kidney disease or medullary sponge kidney. Two patients with biallelic *CYP24A1* variants underwent parathyroidectomy for suspected primary hyperparathyroidism (PHPT), without phenotype alleviation postoperatively.

We furthermore identified three patients with primary hyperoxaluria (PH) in our cohort. The diagnosis was already established in the two patients with primary hyperoxaluria type 1 (PH1) and primary hyperoxaluria type 2 (PH2), respectively. The diagnosis was previously unknown in the patient with primary hyperoxaluria type 3 (PH3) due to a homozygous pathogenic *HOGA1* variant. This patient had his first kidney stone episode in his 30s. Metabolic work-up in his late 60s showed CKD stage 2 and a urine oxalate in the upper normal range on free-choice diet, but a strong increase after an instructed one-week diet low in calcium and sodium.

No Mendelian form of nephrolithiasis was detected in the 114 NKSF included in this study (Fig. 1, Fig. 2, Supplementary Table 5).

### LP/P variants predisposing to nephrolithiasis

The most common identified genetic predisposition to nephrolithiasis was renal phosphate wasting due to monoallelic variants in the genes encoding the renal sodium/phosphate co- transporters NaPi-2a and NaPi-2c, *SLC34A1* (n=15)*, SLC34A3* (n=18) or in the gene *SLC9A3R1,* encoding the regulatory interaction protein for NaPi-2a/2c, NHERF, (n=7). LP/P variants in *SLC34A1*, *SLC34A3*, but not in *SLC9A3R1,* were enriched in KSF compared with controls (NKSF and gnomAD, Supplementary Table 5). KSF with LP/P variants in *SLC34A1/A3* or *SLC9A3R1* had similar plasma phosphate and TmP/GFR compared to KSF without genetic diagnosis, but urine calcium in individuals with LP/P variants in *SLC34A3* was higher compared to KSF without genetic diagnosis (Supplemental Figure 4).

We also identified eight patients carrying the same previously described monoallelic LP/P variant (c.458A>G, p.Asn153Ser) in *CLDN16*, encoding the tight junction protein claudin-16. Biallelic pathogenic variants in *CLDN16* cause familial hypomagnesemia with hypercalciuria and nephrocalcinosis (FHHNC)^33^, and an increased prevalence of nephrolithiasis has been observed in monoallelic carriers of pathogenic *CLDN16* variants ^30,34^. Individuals with the monoallelic *CLDN16* variant presented with hypercalciuria and calcium oxalate stones but normal renal function and plasma magnesium. The identified *CLDN16* variant was more prevalent in KSF vs. NKSF and vs. all LP/P-variants in *CLDN16* in gnomAD-NFE, respectively (Supplementary Table 5).

Additionally, eight patients carried monoallelic LP/P variants in *CYP24A1,* and the respective variants were five to eight times more prevalent in KSF vs. NKSF and vs. gnomAD-NFE, respectively (Supplementary Table 5). Overall (i.e., considering the prevalence of all variants), *CYP24A1* variants were not enriched in our cohort compared to gnomAD-NFE (Supplementary Table 5). KSF with mono- and biallelic *CYP24A1* variants both displayed hypercalciuria, but hypercalcemia and/or suppressed parathyroid hormone (PTH) or nephrocalcinosis were only present in KSF with biallelic variants. Urine calcium was higher in KSF with biallelic variants, but KSF with monoallelic variants had a higher number of past stone events (Supplemental Fig. 5).

We furthermore identified six patients with monoallelic LP/P variants in *SLC7A9*. Risk variants were either previously described to cause autosomal-dominant cystinuria (c.544G>A p.(Ala182Thr)) ^35,36^ ^37,38^, or enriched in KSF vs NKSF and gnomAD (c.313G>A p.(Gly105Arg)). In contrast to biallelic variants, the six KSF with monoallelic *SLC7A9* variants presented with calcium-containing kidney stones without cystine content, similar to previously reported cases ^39-41^. KSF with monoallelic *SLC7A9* variants had elevated urine cystine, albeit lower levels compared to KSF with biallelic variants in *SLC3A1*/*SLC7A9*. No significant differences in stone number or age at first stone event were detected between the two groups of patients (Supplemental Fig. 5). In seven individuals with cystine-containing kidney stones and two individuals with increased urine cystine, no diagnostic variants in *SLC3A1* or *SLC7A9* could be identified (Supplementary Table 6).

Four KSF had LP/P variants in the gene encoding the calcium-sensing receptor, *CASR*, which is associated with familial hypocalciuric hypercalcaemia type 1 (FHH1), or autosomal-dominant hypocalcemia. The frequency of *CASR* LP/P variants in KSF was significantly higher than LP/P variants in all gnomAD-NFE participants (Supplementary Table 5). The phenotype of these patients was variable: two patients presented with hypercalciuria but normal plasma calcium, phosphate and PTH. One patient had hypercalcemia with a low-normal PTH and low urine calcium. Another patient presented with elevated PTH, hypercalcemia, pronounced hypophosphatemia, hypercalciuria and calcium oxalate dihydrate stones, compatible with the diagnosis of PHPT. Parathyroidectomy led to a complete normalization of the phenotype.

In the 114 NKSF included in this study, exome sequencing revealed a LP/P variant in two individuals, corresponding to a prevalence of variants predisposing to nephrolithiasis of 1.8 % (compared with 8.13% for KSF) (Fig. 1, Fig. 2, Supplementary Table 5).

## DISCUSSION

In this exome-sequencing study in a large, unselected European cohort of 787 adult KSF, we detected a Mendelian kidney stone disease in 23 patients (2.9 %) using stringent diagnostic criteria. This diagnostic yield is significantly lower compared to previous studies conducted in selected groups of KSF ^10,17,42^. Yet the fraction of additional individuals (8.13%) with LP/P variants predisposing to nephrolithiasis is substantial, especially when considering the broad inclusion criteria, the high prevalence of the disease and the potential rate of false-negatives due to stringent molecular genetic diagnosis criteria and technical limitations of exome sequencing, such as inability to detect deep intronic or difficulty in reliably calling copy-number variants. In fact, if Mendelian disease and variants predisposing to nephrolithiasis are combined, the overall diagnostic yield is 11 % (87 of 787 individuals), comparable to neurometabolic disorders or cancer, where exome sequencing is routinely used ^16,43,44^. Interestingly, the diagnostic yield was not different between first time and recurrent stone formers (Supplementary Table 3) and did not differ when stratified by sex (Table 3, Supplementary Table 4). In NKSF, no Mendelian kidney stone disease was detected, and LP/P variants in nephrolithiasis genes were significantly less prevalent compared to KSF (1.8 % vs. 8.1 %).

Cystinuria due to biallelic diagnostic variants in *SLC3A1* or *SLC7A9* was the most common Mendelian disease in our cohort (57 %; n=13 of 23 individuals with Mendelian disease), and an additional six individuals carried monoallelic LP/P variants in *SLC7A9*. We confirmed a high prevalence of CKD in KSF with cystinuria ^45^. Yet, this applied only to KSF with biallelic diagnostic variants. We were unable to provide a genetic diagnosis in nine KSF with a cystinuria phenotype, including seven individuals with cystine-containing kidney stones and two individuals with elevated urinary cystine excretion (Supplementary Table 5). While this could be explained at least in part by technical issues, likely additional, yet to be discovered genes, are associated with the clinical phenotype of cystinuria.

Of the LP/P variants detected predisposing to nephrolithiasis, by far the most common (63 %; n=40 of 64) were variants in *SLC34A1/3* or *SLC9A3R1*, highlighting the pathophysiological importance of renal phosphate loss in increasing risk for kidney stone formation ^12,46^. However, the phenotype of KSF with LP/P variants in these genes on a free choice diet was very subtle and indistinguishable from KSF without predisposing variants. Future studies need to determine if the phenotype of KSF with LP/P variants in these genes can be unmasked by dietary interventions (e.g., by a low phosphate diet) for diagnostic purposes and mitigated for prevention of future stone events (e.g., by phosphate supplementation).

We only detected one KSF with a monoallelic LP/P variant in *ADCY10,* indicating that variants in *ADCY10* are not contributing significantly to the overall nephrolithiasis risk, at least in individuals of European descent ^39,47^. Further, we identified four patients with *CASR* LP/P variants in our cohort with very variable phenotypes. Atypical characteristics such as chondrocalcinosis or kidney stones have been described in patients with FHH1 but complicate the separation from primary hyperparathyroidism (PHPT) ^48-50^. Yet, separation is of critical importance: parathyroidectomy is considered as a first-line treatment in PHPT but surgery is ineffective in FHH1. Additional complexity arises from the fact that PHPT and FHH1 can co- exist, as observed in patient# 6023 ^51,52^.

Genetic diagnoses in KSF can have important prognostic implications and therapeutic consequences. However, while the pathophysiology is well delineated, the effectiveness of therapeutic interventions has not been studied systematically in most forms of Mendelian nephrolithiasis. The widespread use of genetic testing will facilitate the inclusion of KSF with Mendelian disease and LP/P variants predisposing to nephrolithiasis in prospective registries and clinical trials to study disease evolution and evaluate tailored therapeutic interventions. Currently, KSF are only subjected to genetic testing if a specific disease is strongly suspected.

While clinical signs suggesting an inherited disorder have been proposed ^53^, Mendelian disease was missed or misclassified in five of 23 (22%) KSF in our cohort, despite individual review of each case by a kidney stone expert. The situation is even far more difficult in KSF with LP/P variants predisposing to nephrolithiasis: the associated phenotypes were discrete and variable with a large phenotype overlap with KSF without LP/P variants. Yet, the low prevalence of LP/P variants detected in NKSF suggests a rather high penetrance of LP/P variants predisposing to nephrolithiasis in adults, with the limitation of a small NKSF sample size. Thus, together, these results preclude the definition of clear phenotypic criteria to prioritize genetic testing in adult KSF. In addition, LP/P variants present with their own challenges in clinics and considerations for clinical reporting of LP/P variants and risk alleles have recently been suggested ^14^.

Strengths of our study include the lack of patient pre-selection with broad inclusion criteria, the large sample size, availability of ethnically matched non-stone forming controls and the detailed phenotype. Our study also has limitations, such as the inability to detect deep intronic and copy-number variants, lack of family recruitment for segregation analyses, overrepresentation of men and a limited ethnic diversity.

In conclusion, our study shows that Mendelian disease is far less common in adult KSF than previously suggested, but simultaneously reveals a high prevalence of LP/P variants predisposing to nephrolithiasis in adult KSF. Our study highlights the potential of genetic testing in unselected adult KSF for accurate assessment of Mendelian disease and identification of variants predisposing to nephrolithiasis to direct patients to tailored therapies and clinical trials.

## DISCLOSURES

The authors declare no competing interests.

## FUNDING

MAA was supported by a grant for protected research time by the InselGruppe. EGO was supported by Postdoc MobilityStipendien of the Swiss National Science Foundation (grants # P2ZHP3_195181 and P500PB_206851), and Kidney Research UK ((grants # Paed_RP_001_20180925). DGF was supported by the Swiss National Science Foundation (grants # 31003A_172974 and 33IC30_166785/1) and the Swiss National Centre of Competence in Research Kidney.CH. JAS is supported by the Northern Counties Kidney Research Fund (22/01), Kidney Research UK (Paed_RP_001_20180925), LifeArc, and the Medical Research Council (MR/Y007808/1).

## Data Availability

All data supporting the findings of the present study are available upon reasonable request to the corresponding author.

## ACKNOWLEDGEMENTS

We wish to thank all the patients and staff from all units participating in the study. We also wish to thank Hans Anderegg for technical expertise in database development.

## AUTHOR CONTRIBUTIONS

DGF, AS and JAS conceptualized the study. DGF and MAA acquired financial support. MAA, EGO and DGF designed data analysis plans. MAA, EGO and MB performed data analysis. MAA, EGO, MB, AS and DGF performed data analysis and data interpretation. MAA wrote the first draft of the manuscript. All authors contributed to discussion and editing of text and approved the final version of the manuscript.

## DATA PRIVACY PROTECTION STATEMENT

Any participant identifiers (PIDs) included were solely used for the BKSR and not known to anyone (e.g., hospital staff, patients or participants themselves) outside the research group. Therefore, they cannot be used to identify individuals.

## DATA SHARING STATEMENT

The data supporting the findings of this study are available from the corresponding author upon request. Stratified genetic data will be accessible upon publication at European Genome-Phenome Archive (EGA) (https://ega-archive.org/).

## SUPPLEMENTARY MATERIAL

**Figure S1:** Stratification pathway used for whole exome sequencing (WES) data in the Bern Kidney Stone Registry (BKSR)

**Figure S2:** 24h urinary risk factors stratified by WES results.

**Figure S3.** Age of first visit and age of first stone event stratified by type of genetic diagnosis (Mendelian vs LP/P predisposing variants).

**Figure S4.** Genotype–phenotype correlations for LP/P phosphate transporter variants.

**Figure S5.** Genotype–phenotype correlations (Cystinuria and *CYP24A1*).

**Table S1.** Genotype and phenotype of patients with LP/P variants predisposing to nephrolithiasis.

**Table S2:** List of likely pathogenic (LP) or pathogenic (P) variants in BKSR

**Table S3.** Characteristics of stone formers stratified by stone events number

**Table S4.** Characteristics of stone formers with diagnostic variants (Mendelian and LP/P predisposing variants) stratified by sex

**Table S5:** Case/control prevalence (enrichment) calculation for LP/P variants

**Table S6.** Patients with cystine stones, clinical cystinuria or elevated urinary cystine but without diagnostic variants in cystinuria genes (*SLC3A1/SLC7A9*)

## Notes

### Competing Interest Statement

The authors have declared no competing interest.

### Author Declarations

The ethical committee of the Kanton of Bern gave ethical approval for this work.

### Summary of Updates

Genetic disease has been separated into clearly Mendelian vs genetic predisposition to nephrolithiasis. This results in revised Fig.1,2, Table 2,3, revised SI and supplementary tables.

## REFERENCES

1. Chewcharat A, Curhan G. Trends in the prevalence of kidney stones in the United States from 2007 to 2016. Urolithiasis. 2021;49(1):27–39.

2. Ferraro PM, Curhan GC, D’Addessi A, Gambaro G. Risk of recurrence of idiopathic calcium kidney stones: analysis of data from the literature. J Nephrol. 2017;30(2):227–233.

3. New F, Somani BK. A Complete World Literature Review of Quality of Life (QOL) in Patients with Kidney Stone Disease (KSD). Curr Urol Rep. 2016;17(12):88.

4. Saigal CS, Joyce G, Timilsina AR. Direct and indirect costs of nephrolithiasis in an employed population: opportunity for disease management? Kidney Int. 2005;68(4):1808–1814.

5. Pearle MS, Goldfarb DS, Assimos DG, et al. Medical management of kidney stones: AUA guideline. The Journal of urology. 2014;192(2):316–324.

6. Dhayat NA, Bonny O, Roth B, et al. Hydrochlorothiazide and Prevention of Kidney-Stone Recurrence. N Engl J Med. 2023;388(9):781–791.

7. Curhan GC, Willett WC, Rimm EB, Stampfer MJ. Family history and risk of kidney stones. J Am Soc Nephrol. 1997;8(10):1568–1573.

8. Goldfarb DS, Fischer ME, Keich Y, Goldberg J. A twin study of genetic and dietary influences on nephrolithiasis: a report from the Vietnam Era Twin (VET) Registry. Kidney Int. 2005;67(3):1053–1061.

9. Edvardsson VO, Palsson R, Indridason OS, Thorvaldsson S, Stefansson K. Familiality of kidney stone disease in Iceland. Scand J Urol Nephrol. 2009;43(5):420–424.

10. Halbritter J, Baum M, Hynes AM, et al. Fourteen monogenic genes account for 15% of nephrolithiasis/nephrocalcinosis. J Am Soc Nephrol. 2015;26(3):543–551.

11. Sadeghi-Alavijeh O, Chan MMY, Moochhala SH, et al. Rare variants in the sodium-dependent phosphate transporter gene SLC34A3 explain missing heritability of urinary stone disease. Kidney Int. 2023.

12. Nwachukwu C, Singh G, Moore B, Strande NT, Bucaloiu ID, Chang AR. Risk of Nephrolithiasis in Adults Heterozygous for SLC34A3 Ser192Leu in an Unselected Health System Cohort. J Am Soc Nephrol. 2023.

13. Halbritter J. Urinary stone disease: closing the heritability gap by challenging conventional Mendelian inheritance. Kidney International. 2023;104(5):882–885.

14. Lebo M, Steeves M, Benson K, et al. O31: Risk allele evidence curation, classification, and reporting: Recommendations from the ClinGen Low Penetrance/Risk Allele Working Group*. Genetics in Medicine Open. 2023;1(1).

15. Groopman EE, Marasa M, Cameron-Christie S, et al. Diagnostic Utility of Exome Sequencing for Kidney Disease. N Engl J Med. 2019;380(2):142–151.

16. Tarailo-Graovac M, Shyr C, Ross CJ, et al. Exome Sequencing and the Management of Neurometabolic Disorders. N Engl J Med. 2016;374(23):2246–2255.

17. Daga A, Majmundar AJ, Braun DA, et al. Whole exome sequencing frequently detects a monogenic cause in early onset nephrolithiasis and nephrocalcinosis. Kidney Int. 2018;93(1):204–213.

18. Braun DA, Lawson JA, Gee HY, et al. Prevalence of Monogenic Causes in Pediatric Patients with Nephrolithiasis or Nephrocalcinosis. Clin J Am Soc Nephrol. 2016;11(4):664–672.

19. Amar A, Majmundar AJ, Ullah I, et al. Gene panel sequencing identifies a likely monogenic cause in 7% of 235 Pakistani families with nephrolithiasis. Hum Genet. 2019;138(3):211–219.

20. Dhayat NA, Schaller A, Albano G, et al. The Vacuolar H+-ATPase B1 Subunit Polymorphism p.E161K Associates with Impaired Urinary Acidification in Recurrent Stone Formers. J Am Soc Nephrol. 2016;27(5):1544-1554.

21. Dhayat NA, Luthi D, Schneider L, Mattmann C, Vogt B, Fuster DG. Distinct phenotype of kidney stone formers with renal phosphate leak. Nephrol Dial Transplant. 2019;34(1):129–137.

22. Dhayat NA, Schneider L, Popp AW, et al. Predictors of Bone Mineral Density in Kidney Stone Formers. Kidney Int Rep. 2022;7(3):558–567.

23. Fuster DG, Morard GA, Schneider L, et al. Association of urinary sex steroid hormones with urinary calcium, oxalate and citrate excretion in kidney stone formers. Nephrol Dial Transplant. 2022;37(2):335–348.

24. DePristo MA, Banks E, Poplin R, et al. A framework for variation discovery and genotyping using next-generation DNA sequencing data. Nat Genet. 2011;43(5):491–498.

25. Poplin R, Ruano-Rubio V, DePristo MA, et al. Scaling accurate genetic variant discovery to tens of thousands of samples. bioRxiv. 2018:201178.

26. McLaren W, Gil L, Hunt SE, et al. The Ensembl Variant Effect Predictor. Genome Biol. 2016;17(1):122.

27. Karczewski KJ, Francioli LC, Tiao G, et al. The mutational constraint spectrum quantified from variation in 141,456 humans. Nature. 2020;581(7809):434-443.

28. Policastro LJ, Saggi SJ, Goldfarb DS, Weiss JP. Personalized Intervention in Monogenic Stone Formers. J Urol. 2018;199(3):623–632.

29. Richards S, Aziz N, Bale S, et al. Standards and guidelines for the interpretation of sequence variants: a joint consensus recommendation of the American College of Medical Genetics and Genomics and the Association for Molecular Pathology. Genet Med. 2015;17(5):405–424.

30. Weber S, Schneider L, Peters M, et al. Novel paracellin-1 mutations in 25 families with familial hypomagnesemia with hypercalciuria and nephrocalcinosis. J Am Soc Nephrol. 2001;12(9):1872–1881.

31. Tebben PJ, Milliner DS, Horst RL, et al. Hypercalcemia, hypercalciuria, and elevated calcitriol concentrations with autosomal dominant transmission due to CYP24A1 mutations: effects of ketoconazole therapy. J Clin Endocrinol Metab. 2012;97(3):E423–427.

32. Hanna C, Potretzke TA, Cogal AG, et al. High Prevalence of Kidney Cysts in Patients With CYP24A1 Deficiency. Kidney Int Rep. 2021;6(7):1895–1903.

33. Simon DB, Lu Y, Choate KA, et al. Paracellin-1, a renal tight junction protein required for paracellular Mg2+ resorption. Science. 1999;285(5424):103-106.

34. Praga M, Vara J, Gonzalez-Parra E, et al. Familial hypomagnesemia with hypercalciuria and nephrocalcinosis. Kidney Int. 1995;47(5):1419–1425.

35. Font-Llitjos M, Jimenez-Vidal M, Bisceglia L, et al. New insights into cystinuria: 40 new mutations, genotype-phenotype correlation, and digenic inheritance causing partial phenotype. J Med Genet. 2005;42(1):58–68.

36. Gaildrat P, Lebbah S, Tebani A, et al. Clinical and molecular characterization of cystinuria in a French cohort: relevance of assessing large-scale rearrangements and splicing variants. Mol Genet Genomic Med. 2017;5(4):373–389.

37. Font MA, Feliubadalo L, Estivill X, et al. Functional analysis of mutations in SLC7A9, and genotype-phenotype correlation in non-Type I cystinuria. Hum Mol Genet. 2001;10(4):305–316.

38. Reig N, Chillaron J, Bartoccioni P, et al. The light subunit of system b(o,+) is fully functional in the absence of the heavy subunit. EMBO J. 2002;21(18):4906–4914.

39. Halbritter J, Baum M, Hynes AM, et al. Fourteen Monogenic Genes Account for 15% of Nephrolithiasis/Nephrocalcinosis. J Am Soc Nephrol. 2014.

40. Elkoushy MA, Andonian S. Characterization of patients with heterozygous cystinuria. Urology. 2012;80(4):795–799.

41. Cupisti A, Farnesi I, Armillotta N, Francesca F. Staghorn cystine stone in a 72-year-old recurrent calcium stone former. Clin Nephrol. 2012;78(1):76–80.

42. Schonauer R, Scherer L, Nemitz-Kliemchen M, et al. Systematic assessment of monogenic etiology in adult-onset kidney stone formers undergoing urological intervention-evidence for genetic pretest probability. Am J Med Genet C Semin Med Genet. 2022;190(3):279–288.

43. Huang KL, Mashl RJ, Wu Y, et al. Pathogenic Germline Variants in 10,389 Adult Cancers. Cell. 2018;173(2):355–370 e314.

44. Zhang J, Walsh MF, Wu G, et al. Germline Mutations in Predisposition Genes in Pediatric Cancer. N Engl J Med. 2015;373(24):2336–2346.

45. Prot-Bertoye C, Lebbah S, Daudon M, et al. CKD and Its Risk Factors among Patients with Cystinuria. Clin J Am Soc Nephrol. 2015;10(5):842–851.

46. Sadeghi-Alavijeh O, Chan MMY, Moochhala SH, et al. Rare variants in in the sodium-dependent phosphate transporter gene SLC34A3 explain missing heritability of urinary stone disease. Kidney International. 2023.

47. Reed BY, Gitomer WL, Heller HJ, et al. Identification and characterization of a gene with base substitutions associated with the absorptive hypercalciuria phenotype and low spinal bone density. J Clin Endocrinol Metab. 2002;87(4):1476–1485.

48. Mouly C, Vargas-Poussou R, Lienhardt A, et al. Clinical characteristics of familial hypocalciuric hypercalcaemia type 1: A multicentre study of 77 adult patients. Clin Endocrinol (Oxf). 2020;93(3):248–260.

49. Volpe A, Guerriero A, Marchetta A, Caramaschi P, Furlani L. Familial hypocalciuric hypercalcemia revealed by chondrocalcinosis. Joint Bone Spine. 2009;76(6):708–710.

50. Stratta P, Merlotti G, Musetti C, et al. Calcium-sensing-related gene mutations in hypercalcaemic hypocalciuric patients as differential diagnosis from primary hyperparathyroidism: detection of two novel inactivating mutations in an Italian population. Nephrol Dial Transplant. 2014;29(10):1902–1909.

51. Eldeiry LS, Ruan DT, Brown EM, Gaglia JL, Garber JR. Primary hyperparathyroidism and familial hypocalciuric hypercalcemia: relationships and clinical implications. Endocr Pract. 2012;18(3):412–417.

52. Egan AM, Ryan J, Aziz MA, O’Dwyer TP, Byrne MM. Primary hyperparathyroidism in a patient with familial hypocalciuric hypercalcaemia due to a novel mutation in the calcium-sensing receptor gene. J Bone Miner Metab. 2013;31(4):477–480.

53. Ferraro PM, D’Addessi A, Gambaro G. When to suspect a genetic disorder in a patient with renal stones, and why. Nephrol Dial Transplant. 2013;28(4):811–820.

