## Supplemental Methods, Figures and Tables for "Prevalence and characteristics of genetic disease in adult kidney stone formers"

### SUPPLEMENTAL INFORMATION (SI)

### SUPPLEMENTAL METHODS

#### Study cohort and phenotypic analysis performed in BKSR

All kidney stone formers (KSF), including first-time stone formers, referred for metabolic work-up to at the Department of Nephrology and Hypertension, Bern University Hospital, Bern, Switzerland are invited to participate in the BKSR. The BKSR adheres to the Declaration of Helsinki and was approved by the ethical committee of the Kanton of Bern (approval # BE 95/06). Inclusion criteria for the BKSR are i) written informed consent, ii) age $\geq$ 18 years, and iii) $\geq$ 1 past kidney stone episode [1-4]. There were no pre-specified exclusion criteria. BKSR participants underwent a detailed metabolic work-up under free choice diet and thereafter 1 week of instructed low–calcium (15–20 mmol/d) and low–sodium (100 mmol/d) diet. Blood and urine analyses were performed at the Central Laboratory of the Bern University Hospital. Kidney stone composition analysis was performed at the Central Laboratory of the Bern University Hospital by Fourier transform infrared spectroscopy [5].

Non-stone forming individuals (NKSF) were recruited internally and consist of volunteers and patients seen for evaluation of arterial hypertension. Inclusion criteria for NKSF were i) written informed consent, ii) age $\geq$ 18 years, and iii) no history of past kidney stone episodes and no evidence of asymptomatic nephrolithiasis or nephrocalcinosis on ultrasound at the time of enrolment. All NKSF underwent a metabolic work-up that was similar to BKSR participants (Fig.1)

#### Detailed information on preparation and analysis of genetic data

Genomic DNA was isolated from peripheral blood leucocytes (NucleoSpin, Macherey-Nagel, Switzerland). Exomes were captured using the Agilent SureSelect Human All Exon v6-Kit (Santa Clara, CA), and high-throughput sequencing was performed on a HiSeq X-Ten platform

(Illumina, San Diego, CA). Reads were aligned to the human reference genome (Genome Reference Consortium build 38 (GRCh38)/human genome 38 (hg38) with Burrows-Wheeler Aligner 0.7.17-r1188 (BWA) [6]. BAM files were generated and sorted by SAMtools 1.10 [7]. Duplicates were removed with Picard 2.20.0 (Broad Institute, Cambridge, MA). Variant calling was conducted using Genome analysis Toolkit v3.8 (Broad Institute, Cambridge, MA) to call single nucleotide variants (SNVs) , small insertions and deletions (InDels) . The obtained VCFs were annotated using Ensembl Variant Effect Predictor Release 106 (VEP) [8]. Pooled VCF files of all patients were imported in a dedicated MySQL-database (MariaDB 10.4), and variants filtered for region of interest (exons and splice regions), predicted consequences (protein truncating, mRNA splicing, missense, synonymous, insertion/deletion), genome aggregation database (gnomAD) minor allele frequency (<1% in all populations) [9] to allow analysis of 34 genes previously implicated in Mendelian (monogenic) kidney stone disease according to recommendations of the American College of Medical Genetics and Genomics (ACMG)[10].

**Variant classification criteria used for "LP/P variants" and case/control prevalence (enrichment) calculations**

Monoallelic/heterozygous diagnostic (LP or P) variants in genes with insufficient or debated evidence for monoallelic/autosomal dominant (AD) Mendelian disease but sufficient evidence (e.g., enrichment on stone formers, predicted or experimental evidence for pathogenicity, predicted or experimental evidence for incomplete penetrance) for intermediate effect size from this study and/or from published case/control studies were labeled "LP/P variants predisposing to nephrolithiasis" (e.g., *ADCY10, CASR, SLC4A1, SLC9A3R1)*[11-16]. Similarly, monoallelic LP or P variants in genes with reported biallelic/autosomal recessive (AR) inheritance where evidence for genotype dosage effects has been reported (e.g., for *CLDN16, CYP24A1 SLC7A9,*

*SLC34A1, SLC34A3,*). were labeled "LP/P variants predisposing to nephrolithiasis" [17-22] In both cases, variants are referred to as "LP/P variants" in the manuscript.

Case and control prevalences for monoallelic predicted deleterious (LP/P) variants conferring a potential genetic predisposition for kidney stones (in CLDN16, CASR, CYP24A1, SLC7A9, SLC9A3R1, SLC34A1 SLC34A3) were calculated based on their prevalence in the present study (787 KSF) and the prevalence of selected variants (ClinVar LP/P variants including/and the specific variants identified in this study) in Non-Finnish Europeans in gnomAD. A statistical difference in prevalence (as a potential sign for enrichment) was calculated using the Fishers exact test.

SUPPLEMENTAL FIGURES AND TABLES

**Supplemental Figure 1**

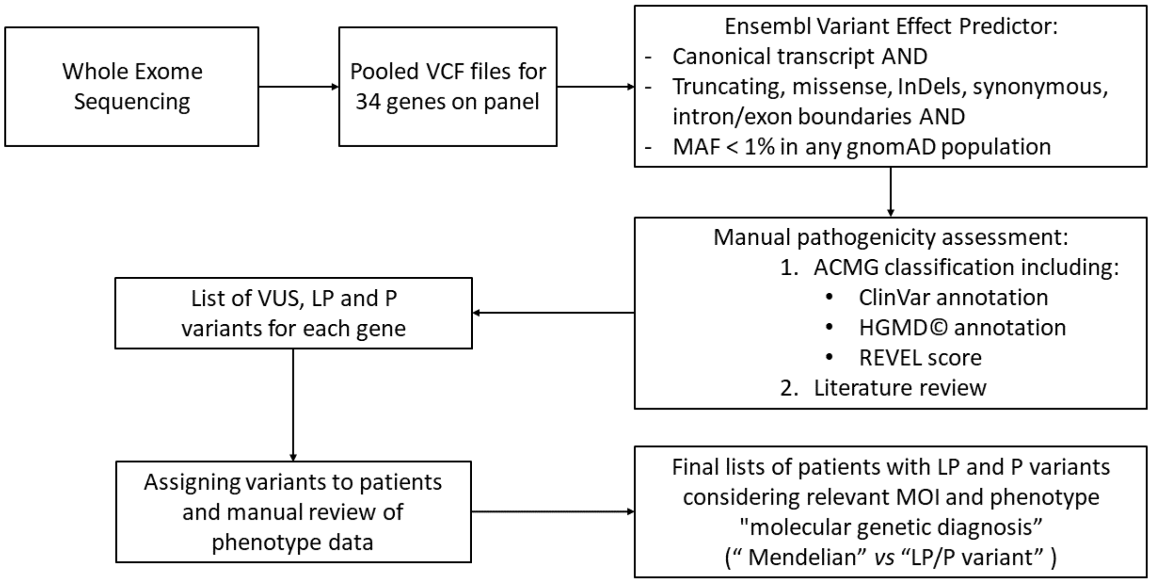


**Stratification pathway used for whole exome sequencing (WES) data in the Bern Kidney Stone Registry (BKSR).** Genetic analysis was performed in a standardized prioritization pathway in 34 known nephrolithiasis genes, followed by interdisciplinary review and classification according to the diagnostic criteria of the American College of Medical Genetics and Genomics (ACMG) with subsequent genotype-phenotype analysis. WES was performed using the Agilent SureSelect Human All Exon v6-Kit on a HiSeq X-10 platform from Illumina©. Only variants classified as likely pathogenic (LP; ACMG class IV) or pathogenic (P; ACMG class V) were defined as diagnostic variants after automated classification using Varsome & Franklin and independent manual review by 3 clinician scientists and 1 geneticist. ACMG: American College of Medical Genetics and Genomics, gnomAD: Genome Aggregation Database, HGMD©: Human Gene Mutation Database, LP: Likely pathogenic, MAF: Minor allele frequency, MOI: Mode of inheritance, P: Pathogenic, REVEL: Rare exome variant ensemble learner, VCF: Variant Call Format, VUS: Variant of unknown significance.

### Supplemental Figure 2

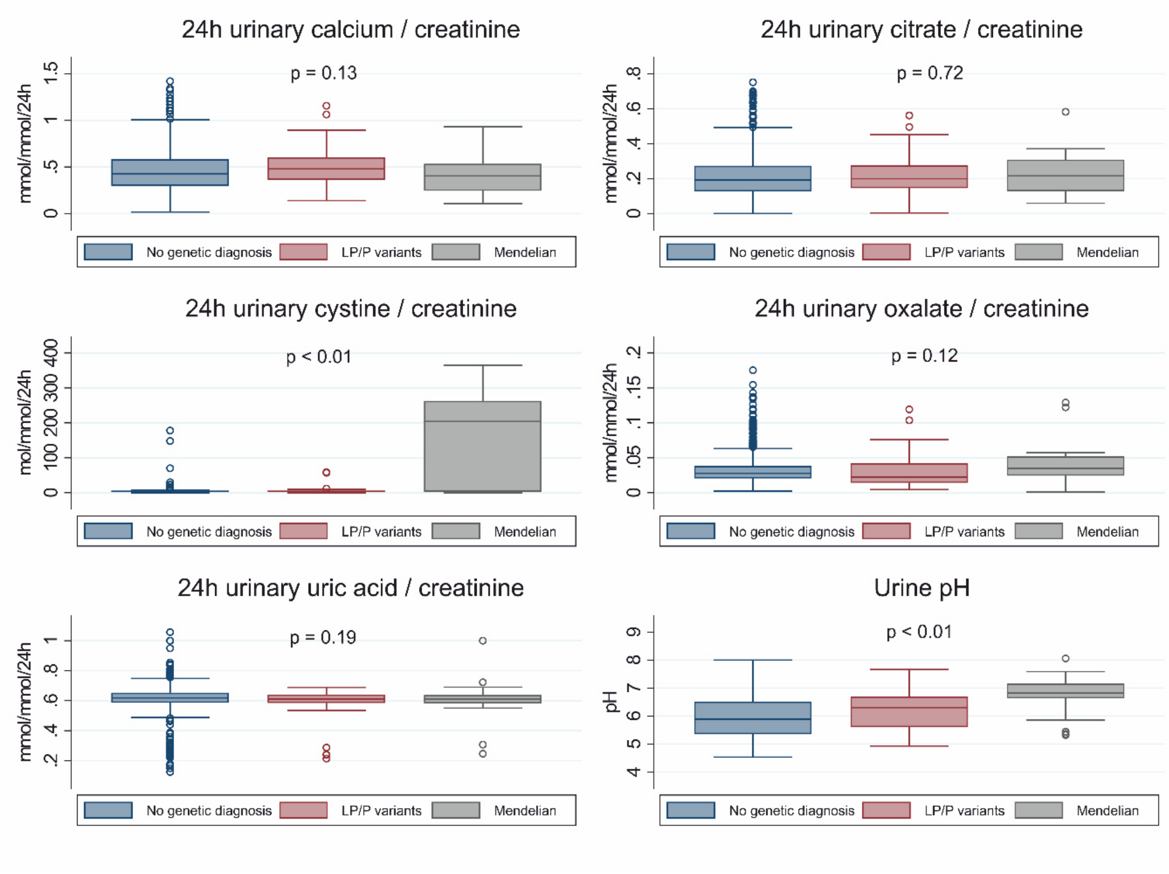


**24h urinary risk factors stratified by WES results.** Boxplots of promotors and inhibitors of stone formation between participants without genetic diagnosis, with Mendelian diagnosis and with LP/P variants predisposing to nephrolithiasis (LP/P variants) in the BKSR. Values reported are creatinine-normalized 24h urines solute excretions, except urine pH. Boxplots show medians with 25th-75th percentiles. Whiskers represent the range of values within 1.5 times the interquartile range from the first and third quartiles. Hollow circles are outliers.

**Supplemental Figure 3**

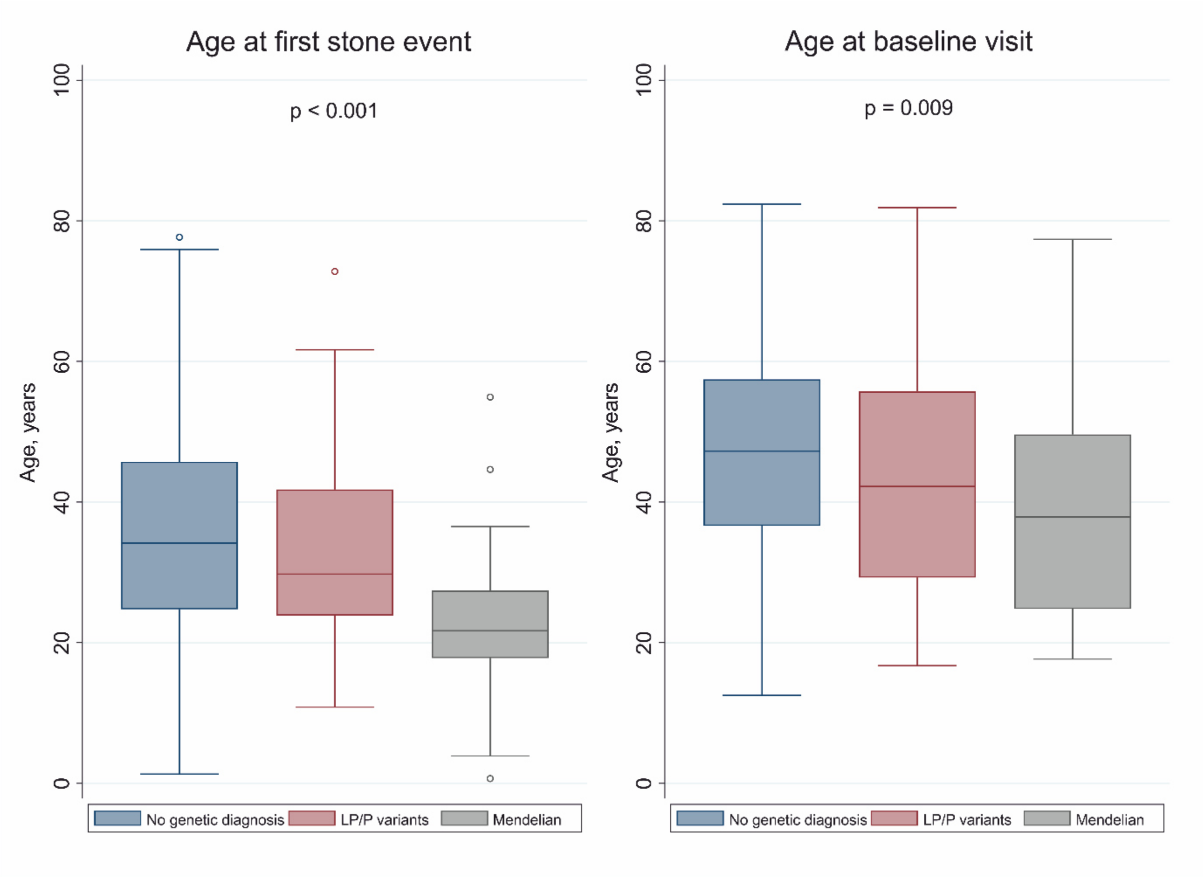


**Age of first visit and age of first stone event stratified by type of genetic diagnosis (Mendelian vs LP/P).** Boxplots of the ages at first visit and first stone event in the BKSR stratified by patients without genetic diagnosis, with Mendelian diagnosis and with LP/P variants predisposing to nephrolithiasis (LP/P variants). Boxplots show medians with 25th-75th percentiles. Whiskers represent the range of values within 1.5 times the interquartile range from the first and third quartiles. Hollow circles are outliers.

**Supplemental Figure 4**

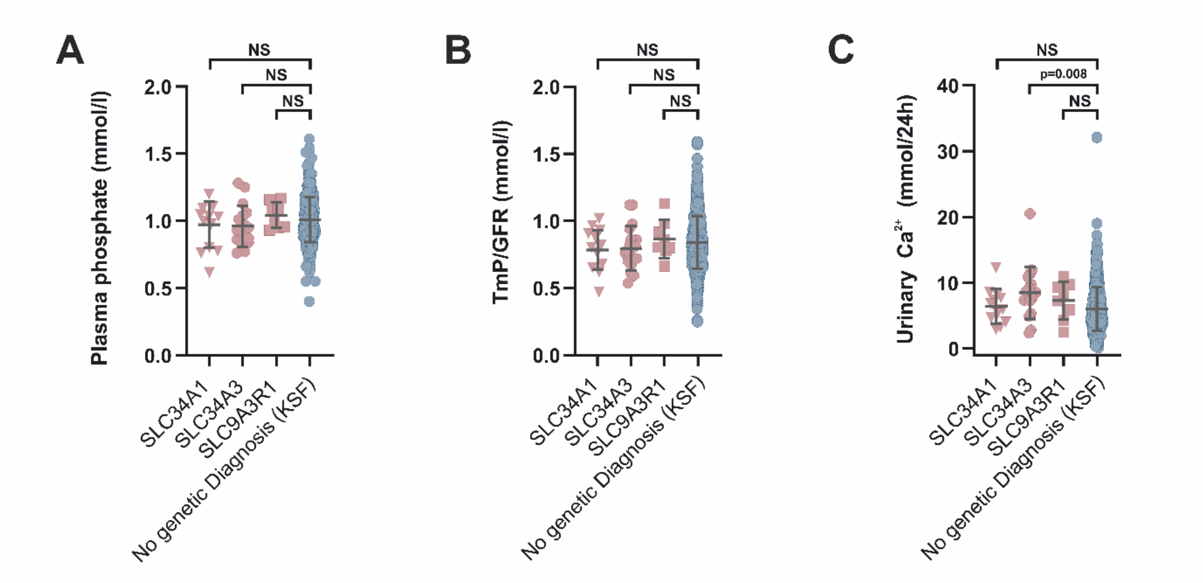


**Genotype–phenotype correlations for monoallelic LP/P phosphate transporter variants.** Plasma phosphate (A), urinary phosphate loss visualized by Tmp/GFR (B), Urine calcium/24h stratified according to genetic diagnosis for urinary phosphate loss (C), Each dot represents combined data from available values/individual. Data are shown as observations/individual with mean ± SD. Statistical analysis: Mann-Whitney U test or Kruskal-Wallis-test, where applicable, were used for non-normally distributed continuous variables.

**Supplemental Figure 5**

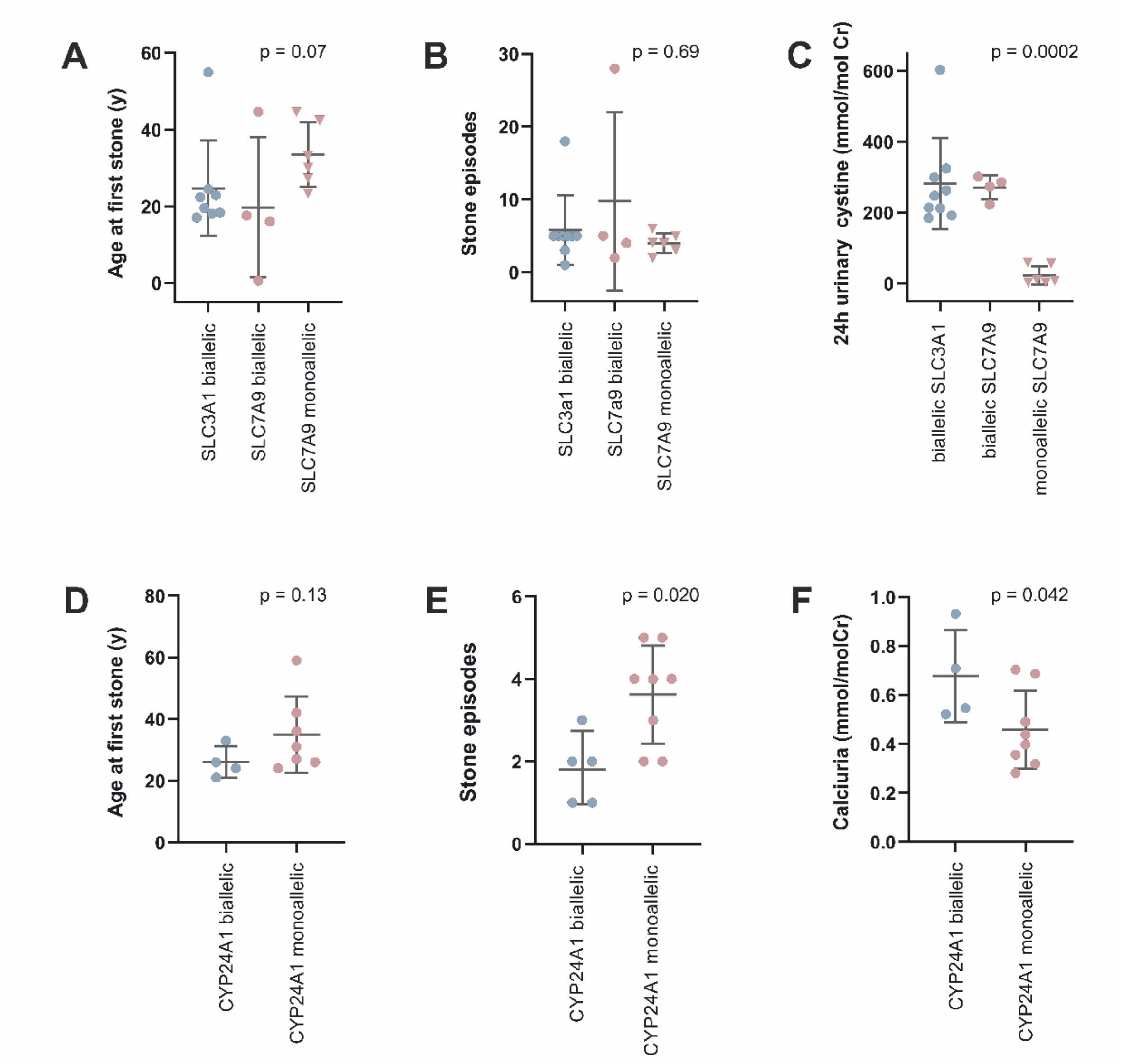


**Genotype–phenotype correlations.** Cystinuria (A-C): Age at 1$^{st}$ stone (A), number of stone episodes (B), Urinary Cystine stratified according to genetic diagnosis for cystinuria (C), *CYP24A1*-variants (D-F): age at 1$^{st}$ stone, number of stone episodes (E), Urinary calcium stratified according to genetic diagnosis (F). Each dot represents combined data from available values/individual. Data are shown as observations/individual with mean ± SD. Statistical analysis: Mann-Whitney U test or Kruskal-Wallis-test, where applicable, were used for non-normally distributed continuous variables.

**Supplementary Table 1.** Genotype and phenotype of patients with LP/P variants predisposing to nephrolithiasis): see attached file "Suppl_Table_1".

**Supplementary Table 2:** List of likely pathogenic (LP) or pathogenic (P) variants (including LP/P variants predisposing to nephrolithiasis) in BKSR: see separate table "Suppl_Table_2".

**Supplementary Table 3 | Characteristics of stone formers stratified by stone events number.** Characteristics are indicated separately for first time and recurrent (> 1 stone event) stone formers. Categorical variables are described by number of participants N (%), continuous variables are described by their mean (SD) or median ($25^{th}$-$75^{th}$ percentile). eGFR, estimated glomerular filtration rate; BSA, body surface area; PTH, parathyroid hormone; SD, standard deviation.

| **Characteristics** | **First time stone formers (N=134)** | **Recurrent stone formers (N=614)** | ***p*-values** |
|---|---|---|---|
| Age, years | 38.86 (15.88) | 48.31 (13.48) | <0.001 |
| Males | 85 (63.4%) | 458 (74.6%) | 0.007 |
| Body mass index, kg/m$^2$ | 25.58 (5.40) | 27.08 (4.94) | 0.002 |
| Hypertension | 38 (29.5%) | 216 (37.6%) | 0.086 |
| Diabetes | 3 (2.3%) | 20 (3.3%) | 0.78 |
| Obesity, BMI > 30 kg/m$^2$ | 23 (17.8%) | 140 (23.0%) | 0.24 |
| Hyperuricemia | 51 (38.3%) | 270 (44.0%) | 0.25 |
| Family history of kidney stone disease | 55 (42.6%) | 262 (43.8%) | 0.85 |
| Kidney stone recurrence, > 1 stone event | 0 (0.0%) | 614 (100.0%) | <0.001 |
| Age at first stone event, years | 36.97 (15.84) | 35.07 (13.37) | 0.16 |
| Total number of stone events | 1.00 (1.00, 1.00) | 3.00 (2.00, 5.00) | <0.001 |
| Mendelian diagnosis | 4 (3.0%) | 19 (3.1%) | 0.97 |
| LP/P variants | 14 (10.4%) | 50 (8.1%) | 0.39 |
| **Blood parameters** | | | |
| Total calcium, mmol/L | 2.36 (0.12) | 2.35 (0.12) | 0.89 |
| Ionized calcium, mmol/L | 1.21 (1.19, 1.24) | 1.21 (1.19, 1.23) | 0.76 |
| Phosphorus, mmol/L | 1.04 (0.16) | 1.00 (0.17) | 0.008 |
| Magnesium, mmol/L | 0.82 (0.06) | 0.83 (0.07) | 0.58 |
| Uric acid, umol/L | 343.35 (369.69) | 325.66 (82.28) | 0.29 |
| Intact PTH, ng/L | 35.00 (27.30, 45.00) | 40.88 (30.50, 53.00) | 0.002 |
| 25-OH Vitamin $D_3$, nmol/L | 40.00 (23.00, 64.00) | 40.36 (27.09, 59.00) | 0.96 |
| eGFR creatinine Equation CKD-EPI 2009, mL/min per 1.73 m$^2$ BSA | 102.77 (88.91, 119.15) | 94.66 (80.34, 107.55) | <0.001 |
| **Urine parameters** | | | |
| Total urine volume, L/24h | 1.83 (1.40, 2.51) | 2.02 (1.49, 2.54) | 0.11 |
| Urine pH | 5.80 (5.32, 6.50) | 5.96 (5.40, 6.61) | 0.25 |
| Urine sodium / creatinine ratio, mmol/mmol/24h | 13.28 (10.76, 16.85) | 13.64 (11.04, 16.99) | 0.61 |
| Urine potassium / creatinine ratio, mmol/mmol/24h | 4.61 (3.61, 5.68) | 4.71 (3.83, 5.88) | 0.49 |
| Urine uric acid / creatinine ratio, mmol/mmol/24h | 0.60 (0.12) | 0.62 (0.10) | 0.052 |
| Urine calcium / creatinine ratio, mmol/mmol/24h | 0.43 (0.31, 0.56) | 0.43 (0.30, 0.60) | 0.62 |
| Urine magnesium / creatinine ratio, mmol/mmol/24h | 0.31 (0.25, 0.38) | 0.30 (0.23, 0.38) | 0.75 |
| Urine citrate / creatinine ratio, mmol/mmol/24h | 0.20 (0.12, 0.27) | 0.19 (0.13, 0.28) | 0.97 |
| Urine phosphate / creatinine ratio, mmol/mmol/24h | 2.15 (0.53) | 2.20 (0.54) | 0.39 |
| Urine oxalate / creatinine ratio, mmol/mmol/24h | 0.03 (0.02, 0.03) | 0.03 (0.02, 0.04) | 0.067 |
| Urine cystine / creatinine ratio, mmol/mol/24h | 4.00 (3.00, 5.00) | 4.00 (3.00, 6.00) | 0.065 |
| **Stone phenotypes** | | | |
| Calcium oxalate dihydrate | 11 (12.8%) | 62 (13.0%) | 1.00 |
| Calcium oxalate monohydrate | 46 (53.5%) | 241 (50.4%) | 0.64 |
| Calcium phosphate | 19 (22.1%) | 111 (23.2%) | 0.89 |
| Uric acid | 2 (2.3%) | 35 (7.3%) | 0.099 |
| Cystine | 4 (4.7%) | 14 (2.9%) | 0.50 |
| Struvite | 3 (3.5%) | 12 (2.5%) | 0.49 |

**Supplementary Table 4 | Characteristics of stone formers with diagnostic variants (Mendelian and LP/P variants predisposing to nephrolithiasis) stratified by sex.** Categorical variables are described by number of participants N (%), continuous variables are described by their mean (SD) or median ($25^{th}$-$75^{th}$ percentile). eGFR, estimated glomerular filtration rate; BSA, body surface area; PTH, parathyroid hormone; SD, standard deviation.

| Characteristics | Females (N=23) | Males (N=64) | *p*-values |
|---|---|---|---|
| Age, years | 40.10 (16.43) | 43.45 (15.64) | 0.39 |
| Body mass index, kg/m$^2$ | 25.43 (5.31) | 26.25 (4.33) | 0.47 |
| Hypertension | 10 (43%) | 22 (40%) | 0.80 |
| Diabetes | 0 (0%) | 1 (2%) | 1.00 |
| Obesity, BMI > 30 kg/m$^2$ | 5 (22%) | 11 (18%) | 0.76 |
| Hyperuricemia | 6 (26%) | 34 (53%) | 0.030 |
| Family history of kidney stone disease | 9 (41%) | 29 (49%) | 0.62 |
| Kidney stone recurrence, > 1 stone event | 17 (74%) | 50 (82%) | 0.54 |
| Age at first stone event, years | 27.42 (13.73) | 32.79 (13.75) | 0.12 |
| Total number of stone events | 3.00 (1.00, 5.00) | 3.00 (2.00, 4.00) | 0.84 |
| **Blood parameters** | | | |
| Total calcium, mmol/L | 2.36 (0.11) | 2.39 (0.12) | 0.41 |
| Ionized calcium, mmol/L | 1.21 (1.17, 1.23) | 1.22 (1.20, 1.25) | 0.11 |
| Phosphorus, mmol/L | 1.03 (0.15) | 0.98 (0.14) | 0.20 |
| Magnesium, mmol/L | 0.82 (0.08) | 0.83 (0.07) | 0.34 |
| Uric acid, umol/L | 276.25 (78.67) | 347.93 (65.14) | <0.001 |
| Intact PTH, ng/L | 34.00 (25.40, 57.00) | 37.00 (25.00, 45.00) | 0.84 |
| 25-OH Vitamin $D_3$, nmol/L | 39.00 (23.00, 62.80) | 44.00 (29.00, 62.00) | 0.54 |
| eGFR creatinine Equation CKD-EPI 2009, mL/min per 1.73 m$^2$ BSA | 93.65 (79.29, 100.54) | 98.07 (81.16, 108.74) | 0.32 |
| **Urine parameters** | | | |
| Total urine volume, L/24h | 2.93 (2.28, 3.85) | 2.10 (1.36, 2.51) | <0.001 |
| Urine pH | 6.63 (5.76, 6.90) | 6.43 (5.63, 6.84) | 0.62 |
| Urine sodium / creatinine ratio, mmol/mmol/24h | 18.91 (14.85, 21.55) | 12.69 (10.75, 16.34) | <0.001 |
| Urine potassium / creatinine ratio, mmol/mmol/24h | 6.55 (5.41, 7.35) | 4.44 (3.73, 5.56) | <0.001 |
| Urine uric acid / creatinine ratio, mmol/mmol/24h | 0.60 (0.17) | 0.59 (0.09) | 0.93 |
| Urine calcium / creatinine ratio, mmol/mmol/24h | 0.45 (0.40, 0.64) | 0.44 (0.33, 0.55) | 0.38 |
| Urine magnesium / creatinine ratio, mmol/mmol/24h | 0.40 (0.24, 0.47) | 0.30 (0.23, 0.35) | 0.077 |
| Urine citrate / creatinine ratio, mmol/mmol/24h | 0.24 (0.17, 0.33) | 0.19 (0.14, 0.25) | 0.083 |
| Urine phosphate / creatinine ratio, mmol/mmol/24h | 2.08 (0.44) | 2.17 (0.47) | 0.43 |
| Urine oxalate / creatinine ratio, mmol/mmol/24h | 0.04 (0.02, 0.06) | 0.02 (0.01, 0.04) | 0.018 |
| Urine cystine / creatinine ratio, mmol/mol/24h | 8.00 (3.50, 204.50) | 4.00 (3.00, 8.00) | 0.071 |
| **Stone phenotypes** | | | |
| Calcium oxalate dihydrate | 0 (0%) | 7 (16%) | 0.093 |
| Calcium oxalate monohydrate | 5 (28%) | 18 (42%) | 0.39 |
| Calcium phosphate | 4 (22%) | 11 (26%) | 1.00 |
| Uric acid | 0 (0%) | 1 (2%) | 1.00 |
| Cystine | 7 (39%) | 4 (9%) | 0.011 |
| Struvite | 2 (11%) | 1 (2%) | 0.21 |

**Supplementary Table 5: Case/control prevalence (enrichment) calculation for LP/P variants present in KSF from the BKSR vs NKSF form the BKSR and vs gnomAD (NFE):**
BKSR: Bern Kidney Stone Registry, KSF: Kidney stone formers, NKSF: Non kidney stone forming controls, LP: likely pathogenic, P: pathogenic, NFE: non-finnish europeans.

| Gene | LP/P reported in the BKSR | total KSF in BKSR | Prevalence in BKSR | L/P in NKSF | total NKSF | Prevalence in NKSF | ClinVar LP/P variants reported in gnomAD NFE (and the specific LP/P variants reported in the BKSR) | total gnomAD NFE participants (rounded) | Prevalence in gnomAD (NFE) | p-Value (Fishers) for independence |
|---|---|---|---|---|---|---|---|---|---|---|
| CASR | 4 | 787 | 0.51% | 0 | 114 | 0% | 30 | 57000 | 0.05% | **0.0011** |
| CLDN16 het | 8 | 787 | 1.02% | 0 | 114 | 0% | 94 | 57000 | 0.16% | **<0.0001** |
| CYP24A1 het | 8 | 787 | 1.02% | 0 | 114 | 0% | 542 | 57000 | 0.95% | 0.85 |
| SLC34A1 het | 15 | 787 | 1.91% | 0 | 114 | 0% | 541 | 57000 | 0.95% | **0.014** |
| SLC34A3 het | 18 | 787 | 2.29% | 1 | 114 | 0.88% | 262 | 57000 | 0.46% | **< 0,0001** |
| SLC7A9 het | 7 | 787 | 0.89% | 0 | 114 | 0% | 676 | 57000 | 1.19% | 0.62 |

| Gene | Frequency of R153Q variant in the BKSR | total KSF in BKSR | Prevalence in BKSR | L/P in NKSF | total NKSF | Prevalence in NKSF | Frequency of R153Q variant (no LP/P in gnomAD ) | total gnomAD NFE participants | Prevalence of R153Q in gnoMAD (NFE) | p-Value (Fishers) for independence |
|---|---|---|---|---|---|---|---|---|---|---|
| SLC9A3R1 | 7 | 787 | 0.89% | 1 | 115 | 0.88% | 435 | 64,541 | 0.67% | 0.38 |

**Supplementary Table 6: Patients with cystine stones, clinical cystinuria or elevated urinary cystine but without diagnostic or LP/P variants predisposing to nephrolithiasis in cystinuria genes (*SLC3A1/SLC7A9*)**

M: male, F: female, Family history/Cystine Stone: N= no, Y=yes, NL: Nephrolithiasis, NC: Nephrocalcinosis, CKD: chronic kidney disease. PSC: primary sclerosing cholangitis, Stone analysis: CaOx: Calcium oxalate, COM: Calcium oxalate monohydrate, COD: Calcium oxalate dihydrate, CA: Carbonate apatite. Creat: Creatinine, Urinary cystine: Norm: <30 mmol/molCreat)

| **ID** | **Sex** | **Age at first stone (range)** | **Number of past stone events** | **Stone composition analysis (%)** | **Family history** | **Key Phenotype** | **Clinical diagnosis before WES** | **Cystine Stone** | **Cystine Percentage** | **Urinary Cystine** (mmol/mol Creat.) | **Urinary Ornithine** (mmol/mol Creat.) | **Urinary Lysine** (mmol/mol Creat.) | **Urinary Arginine** (mmol/mol Creat.) |
|---|---|---|---|---|---|---|---|---|---|---|---|---|---|
| 5215 | F | 16-20 | 5 | 100 Cystine | N | Cystine-NL, Cystinuria, Hyperoxaluria | Cystinuria, | Y | 100 | 148 | 70 | 305 | |
| 5466 | M | 41-45 | 1 | 80 COD, 20 Cystine | N | COD/Cystine NL, Hypocitratiuria | PSC, chronic inflammatory bowel disease, | Y | 20 | 3 | 0 | 7 | 0 |
| 5475 | M | 26-30 | 1 | 80 COD, 20 Cystine | N | COD/Cystine NL, Hyperuricemia | Idiopathic NL | Y | 20 | 3 | 0 | 10 | 0 |
| 5480 | M | 21-25 | 4 | 80 COD, 20 Cystine | Y | COD/Cystine NL, Osteopenia | Idiopathic NL | Y | 20 | 4 | 0 | 9 | 0 |
| 5494 | M | 31-35 | 7 | 80 COD, 20 COM | Y | CaOx NL, Cystinuria, Osteopenia | Cystinuria without Cystine Stones | N | 0 | 70 | 35 | 344 | 14 |
| 5513 | M | 46-50 | 5 | N/A | Y | borderline Cystinuria | Idiopathic NL | N | N/A | 30 | 8 | 107 | 7 |
| 5527 | M | 11-15 | 5 | 90 Cystine, 10 CA | Y | Cystine-NL, Cystinuria, Hypocitratiuria, Hyperuricemia | Cystinuria | Y | 90 | 178 | 187 | 642 | 227 |
| 5540 | M | 21-25 | 1 | 80 COM, 20 Cystine | N | COD/Cystine NL, Hyperuricemia | Idiopathic NL | Y | 20 | 3 | 0 | 9 | 0 |
| 5833 | M | 11-15 | 5 | 90 Cystine, 10 CA | N | Cystine-NL, Hyperoxaluria, Hyperuricemia | Cystinuria | Y | 90 | N/A | N/A | N/A | N/A |

**Supplementary references:**


1. Dhayat NA, Schaller A, Albano G, et al. The Vacuolar H+-ATPase B1 Subunit Polymorphism p.E161K Associates with Impaired Urinary Acidification in Recurrent Stone Formers. *J Am Soc Nephrol.* 2016;27(5):1544-1554.
2. Dhayat NA, Luthi D, Schneider L, Mattmann C, Vogt B, Fuster DG. Distinct phenotype of kidney stone formers with renal phosphate leak. *Nephrol Dial Transplant.* 2019;34(1):129-137.
3. Dhayat NA, Schneider L, Popp AW, et al. Predictors of Bone Mineral Density in Kidney Stone Formers. *Kidney Int Rep.* 2022;7(3):558-567.
4. Fuster DG, Morard GA, Schneider L, et al. Association of urinary sex steroid hormones with urinary calcium, oxalate and citrate excretion in kidney stone formers. *Nephrol Dial Transplant.* 2022;37(2):335-348.
5. Cloutier J, Villa L, Traxer O, Daudon M. Kidney stone analysis: "Give me your stone, I will tell you who you are!". *World J Urol.* 2015;33(2):157-169.
6. Li H, Durbin R. Fast and accurate short read alignment with Burrows-Wheeler transform. *Bioinformatics.* 2009;25(14):1754-1760.
7. Danecek P, Bonfield JK, Liddle J, et al. Twelve years of SAMtools and BCFtools. *Gigascience.* 2021;10(2).
8. McLaren W, Gil L, Hunt SE, et al. The Ensembl Variant Effect Predictor. *Genome Biol.* 2016;17(1):122.
9. Karczewski KJ, Francioli LC, Tiao G, et al. The mutational constraint spectrum quantified from variation in 141,456 humans. *Nature.* 2020;581(7809):434-443.
10. Richards S, Aziz N, Bale S, et al. Standards and guidelines for the interpretation of sequence variants: a joint consensus recommendation of the American College of Medical Genetics and Genomics and the Association for Molecular Pathology. *Genet Med.* 2015;17(5):405-424.
11. Reed BY, Gitomer WL, Heller HJ, et al. Identification and characterization of a gene with base substitutions associated with the absorptive hypercalciuria phenotype and low spinal bone density. *J Clin Endocrinol Metab.* 2002;87(4):1476-1485.
12. Daga A, Majmundar AJ, Braun DA, et al. Whole exome sequencing frequently detects a monogenic cause in early onset nephrolithiasis and nephrocalcinosis. *Kidney Int.* 2018;93(1):204-213.
13. Karim Z, Gerard B, Bakouh N, et al. NHERF1 mutations and responsiveness of renal parathyroid hormone. *N Engl J Med.* 2008;359(11):1128-1135.
14. Stratta P, Merlotti G, Musetti C, et al. Calcium-sensing-related gene mutations in hypercalcaemic hypocalciuric patients as differential diagnosis from primary hyperparathyroidism: detection of two novel inactivating mutations in an Italian population. *Nephrol Dial Transplant.* 2014;29(10):1902-1909.
15. Raue F, Pichl J, Dorr HG, et al. Activating mutations in the calcium-sensing receptor: genetic and clinical spectrum in 25 patients with autosomal dominant hypocalcaemia - a German survey. *Clin Endocrinol (Oxf).* 2011;75(6):760-765.
16. Ito N, Ihara K, Kamoda T, et al. Autosomal dominant distal renal tubular acidosis caused by a mutation in the anion exchanger 1 gene in a Japanese family. *CEN Case Rep.* 2015;4(2):218-222.
17. Sadeghi-Alavijeh O, Chan MMY, Moochhala SH, et al. Rare variants in the sodium-dependent phosphate transporter gene SLC34A3 explain missing heritability of urinary stone disease. *Kidney Int.* 2023.

18. Nwachukwu C, Singh G, Moore B, Strande NT, Bucaloiu ID, Chang AR. Risk of Nephrolithiasis in Adults Heterozygous for SLC34A3 Ser192Leu in an Unselected Health System Cohort. *J Am Soc Nephrol.* 2023.
19. Weber S, Schneider L, Peters M, et al. Novel paracellin-1 mutations in 25 families with familial hypomagnesemia with hypercalciuria and nephrocalcinosis. *J Am Soc Nephrol.* 2001;12(9):1872-1881.
20. Tebben PJ, Milliner DS, Horst RL, et al. Hypercalcemia, hypercalciuria, and elevated calcitriol concentrations with autosomal dominant transmission due to CYP24A1 mutations: effects of ketoconazole therapy. *J Clin Endocrinol Metab.* 2012;97(3):E423-427.
21. Cools M, Goemaere S, Baetens D, et al. Calcium and bone homeostasis in heterozygous carriers of CYP24A1 mutations: A cross-sectional study. *Bone.* 2015;81:89-96.
22. Carpenter TO. Take another CYP: confirming a novel mechanism for "idiopathic" hypercalcemia. *J Clin Endocrinol Metab.* 2012;97(3):768-771.

**Supplementary Table 1: Genotype and phenotype of patients with monoallelic LP/P variants for KSD**. Monoallelic LP/P variants in genes with AD-inheritance are seen as intermediate effect variants predisposing to KSD. Monoallelic LP/P variants in genes with AR inheritance with evidence of gene dosage effect are seen as predisposing for KSD. Acc.N°: RefSeq accession number, ACMG: American College of Medical Genetics, AF: allele frequency, AR: autosomal recessive, AD: autosomal dominant, allel: alleles, dbSNP: Reference SNP number of variant, het: heterozygous, hom: homozygous, , LP: likely pathogenic, P: pathogenic, tot: total, Novel: mutation detected for the first time in this study/not previously described, NS: nonsense, Family history: N= no, Y=yes, , M: male, F: female, mo: months, NL: Nephrolithiasis, NC: Nephrocalcinosis, dRTA: distal renal tubular acidosis, PHPTH: primary hyperparathyroidism, CKD: chronic kidney disease. Stone analysis: CaOx: Calcium oxalate, COM: Calcium oxalate monohydrate, COD: Calcium oxalate dihydrate, CaP: Calcium phosphate, CA: Carbonate apatite, Brushite: Calcium hydrogen phosphate dihydrate, MAP: Magnesium ammonium phosphate (=Struvite), UA: Uric acid.

| ID | Gene | Inheri-tance | Allelic state | Nucleotide change | Amino acid change | ACMG Class | ACMG criteria | AF gnoMAD (allel/tot/hom) gnoMAD (NFE) | Acc. N° dbSNP | Sex | Age at first stone (range) | Number of past stone events | Stone composition analysis (%) | Family history | Key Phenotype | Clinical diagnosis before WES | Genetic diagnosis after WES |
|---|---|---|---|---|---|---|---|---|---|---|---|---|---|---|---|---|---|
| | **ADCY10 – Idiopathic Hypercalciuria** | | | | | | | | | | | | | | | | |
| 5750 | *ADCY10* | AD | het | c.4477del | p.Leu1493SerfsTer24 | P | PVS1, PM2, PP5 | 92/251.456/0<br>54/113.736/0 | NM_018417.6<br>rs576118131 | M | 16-20 | 2 | 50 UA, 50 UA dihydrate | N | UA NL, DM II<br>Hypercalciuria (5.3)<br>Low urine pH (4.8) | UA NL | Absorptive hypercalciuria |
| | **CASR –Familial hypocalciuric hypercalcemia type 1 (FHH1) / Autosomal Dominant Hypercalciuria Hypocalcemia (ADH)** | | | | | | | | | | | | | | | | |
| 5026 | *CASR* | AD | het | c.206G>A | p.Arg69His | LP | PM2, PM5, PM1supp, PP2, PP3, PP5 | 1/251.300/0<br>0/113.596/0 | NM_000388.4<br>rs193922432 | F | 16-20 | 1 | N/A | N | NL (radiopaque stones)<br>Hypercalcemia (total: 2.7; ionized 1.37)<br>Normal plasma phosphate (0.9)<br>Normal urine calcium (2.9)<br>Normal PTH (19) | Idiopathic NL | CASR-associated KSD |
| 5666 | *CASR* | AD | het | c.1942C>T | p.Arg648Ter | P | PVS1s, PM2, PP5 | 3/251.356/0<br>1/113.760/0 | NM_000388.4<br>rs1185593894 | M | 36-40 | 3 | 90 COM, 10 UA | Y | CaOx NL, osteopenia<br>Normal plasma calcium (2.46)<br>Hypercalciuria (9.35)<br>Normal PTH (27) | Idiopathic NL | CASR-associated KSD |
| 5686 | *CASR* | AD | het | c.269A>C | p.Asn90Thr | LP | PM2, PM1supp, PP2, PP3, PP5 | 2/251.398/0<br>2/113.682/0 | NM_000388.4<br>rs193922439 | M | 26-30 | 2 | 1) 100 COM; 2) 60 COM, 30 COD, 10 CA | N | CaOx NL<br>Normal plasma calcium (2.44)<br>Hypercalciuria (5.88)<br>Normal PTH (25) | Idiopathic NL | CASR-associated KSD |
| 6023 | *CASR* | AD | het | c.848T>C | p.Ile283Thr | LP | PM2, PP3, PP2, PP5m | 26/251.332/0<br>20/113.650/0 | NM_000388.4<br>rs142745096 | M | N/A | N/A | 2) 90 COD, 5 COD, 5 CA | N | CaOx NL, PHPT<br>Hypercalcemia (total: 2.8; ionized 1.38)<br>Hypophosphatemia (0.52)<br>Hypercalciuria (12.2)<br>High PTH (111) | PHPT, s/p parathyroidect-omy | CASR-associated KSD |

| | | | | | | | | | | | | | | | | |
|---|---|---|---|---|---|---|---|---|---|---|---|---|---|---|---|---|
| | **CYP24A1 – (Infantile) HC/NL, 1,25-OH vitamin D-24 hydroxylase deficiency / CYP24A1-associated kidney stone disease** | | | | | | | | | | | | | | | |
| 5028 | *CYP24A1* | AR | het | c.428_430 del | p.Glu143del | P | PM2, BS2, PM4, PP5vs | 133/251.266/1 119/113.708/1 | NM_000782.5 rs777676129 | M | 56-60 | 3 | 1): 50 COD, 25 CA, 25 MAP, 2): 90 Brushite, 10 COD | N | CaOx NL, osteopenia. Normal plasma calcium (total: 2.44; ionized 1.28) Normal PTH (37) Normal 1,25 Vit D (156) Hypercalciuria (11.35) | Idiopathic NL | CYP24A1-associated kidney stone disease |
| | SLC9A3R1 | AR | het | c.458G>A | p.Arg153Gln | LP | PS3s, PM2, BP6supp | 462/251.340/1 381/113.668/2 | NM_004252.5 rs41282065 | | | | | | | | SLC9A3R1-related kidney stone disease |
| 5037 | *CYP24A1* | AR | het | c.1315C>T | p.Arg439Cys | LP | PP3s, PM2, PP5 | 10/251.358/0 8/113.670/0 | NM_000782.5 rs374292194 | M | 31-35 | 2 | 90 COM, 10 COD | N | CaOx NL, osteopenia Normal plasma calcium (total 2.43; ionized 1.23) Normal PTH (34) Normal 25-Vit D (35) Normal 1,25-Vit D (98) Hypercalciuria (5.8) | Idiopathic NL | CYP24A1-associated kidney stone disease |
| 5313 | *CYP24A1* | AR | het | c.1186C>T | p.Arg396Trp | P | PM2, PM5, PP3m, PP5vs | 167/251.228/1 124/113.676/1 | NM_000782.5 rs114368325 | M | N/A | 4 | 80 COM, 20 COD | Y | CaOx NL, osteopenia Normal plasma calcium (total 2.46; ionized 1.25) Normal PTH (28) Low 25-Vit D (30) Normal 1,25-Vit D (153) Normal urine calcium (2.4) | Idiopathic NL | CYP24A1-associated kidney stone disease |
| 5377 | *CYP24A1* | AR | het | c.1226T>C | p.Leu409Ser | LP | PM2, PP5vs | 188/251.066/0 153/113.656/0 | NM_000782.5 rs6068812 | M | 36-40 | 5 | 1) 95 COM/COD, 5 CA 2) 20 COM, 80 UA 2) 20 COD, 80 UA | N | CaOx NL, Normal plasma calcium (total 2.34; ionized 1.23) Normal PTH (31) Low 25-Vit D (12) Normal 1,25-Vit D (51) Hypercalciuria (8.2) | Idiopathic NL | CYP24A1-associated kidney stone disease |
| 5589 | *CYP24A1* | AR | het | c.428_430del | p.Glu143del | P | PM2, BS2, PM4, PP5vs | 133/251.266/1 119/113.708/1 | NM_000782.5 rs777676129 | M | 21-25 | 2 | 70 COM, 30 COD | Y | CaOx NL, Normal plasma calcium (total 2.21; ionized 1.23) Normal PTH (43) Low 25-Vit D (62) Normal 1,25-Vit D (67) Hypercalciuria (5.6) | Idiopathic NL | CYP24A1-associated kidney stone disease |

| | | | | | | | | | | | | | | | |
|---|---|---|---|---|---|---|---|---|---|---|---|---|---|---|---|
| 5711 | *CYP24A1* | AR | het | c.1186C>T | p.Arg396Trp | P | PM2, PM5, PP3m, PP5vs | 167/251.228/1 124/113.676/1 | NM_000782.5 rs114368325 | M | 26-30 | 5 | 100 COM | Y | CaOx NL, Normal plasma calcium (total 2.38; ionized 1.21) Normal PTH (28) Normal 25-Vit D (84) Normal 1,25-Vit D (108) Hypercalciuria (6.9) | Idiopathic NL | CYP24A1-associated kidney stone disease |
| 5752 | *CYP24A1* | AR | het | c.1315C>T | p.Arg439Cys | LP | PP3s, PM2, PP5 | 10/251.358/0 8/113.670/0 | NM_000782.5 rs374292194 | M | 41-45 | 4 | 1) 90 COD, 5 COM, 5 CA 2) 90 COD, 5 COM, 5 CA | Y | CaOx NL, Osteoporosis Normal plasma calcium (total 2.09; ionized 1.19) Normal PTH (36) Normal 25-Vit D (52) Normal 1,25-Vit D (103) Hypercalciuria (5.8) | Idiopathic NL | CYP24A1-associated kidney stone disease |
| 5803 | *CYP24A1* | AR | het | c.1226T>C | p.Leu409Ser | LP | PM2, PP5vs | 188/251.066/0 153/113.656/0 | NM_000782.5 rs6068812 | M | 26-30 | 2 | 60 COD, 20 COM, 20 CA | N | CaOx NL Normal plasma calcium (total 2.29) Normal PTH (31) Normal 25-Vit D (59) Normal 1,25-Vit D (186) Hypercalciuria (8.3) | Idiopathic NL, incomplete dRTA, medullary NC | CYP24A1-associated kidney stone disease |
| | **CLDN16 – Familial hypomagnesemia with hypercalciuria & nephrocalcinosis (FHHNC) / CLDN-16 associated kidney stone disease** | | | | | | | | | | | | | | | | |
| 5001 | *CLDN16* | AR | het | c.458A>G | p.Asn153Ser | LP | PS4, PM1, PP2, BS2, PP5 | 83/251.464/1 61/113.742/0 | NM_006580.4 rs201367228 | M | 41-45 | 4 | N/A | N | NL, Normal plasma calcium (total 2.47; ionized 1.28) Normal plasma magnesium (0.89) Normal PTH (53) Normal 1,25-Vit D (110) Normal urine calcium (4.3) Normal urine magnesium (2.76) | Idiopathic NL | CLDN16-associated kidney stone disease |

| | | | | | | | | | | | | | | | | | |
|---|---|---|---|---|---|---|---|---|---|---|---|---|---|---|---|---|---|
| 5039 | *CLDN16* | AR | het | c.458A>G | p.Asn153Ser | LP | PS4, PM1, PP2, BS2, PP5 | 83/251.464/1 61/113.742/0 | NM_006580.4 rs201367228 | M | 16-20 | 5 | 90 COD, 10 COM | Y | CaOx NL<br>Normal plasma calcium (total 2.37; ionized 1.19)<br>Normal plasma magnesium (0.86)<br>Normal PTH (40)<br>Normal 1,25-Vit D (148)<br>Hypercalciuria (8.4)<br>Normal urine magnesium (4.82) | Idiopathic NL | CLDN16-associated kidney stone disease |
| 5199 | *CLDN16* | AR | het | c.458A>G | p.Asn153Ser | LP | PS4, PM1, PP2, BS2, PP5 | 83/251.464/1 61/113.742/0 | NM_006580.4 rs201367228 | M | 36-40 | 3 | 80 COM, 20 COD | Y | CaOx NL<br>Normal plasma calcium (total 2.29; ionized 1.2)<br>Normal PTH (49)<br>Low 25-Vit D (10)<br>Low 1,25-Vit D (45)<br>Normal urine calcium (3.7) | Idiopathic NL | CLDN16-associated kidney stone disease |
| 5250 | *CLDN16* | AR | het | c.458A>G | p.Asn153Ser | LP | PS4, PM1, PP2, BS2, PP5 | 83/251.464/1 61/113.742/0 | NM_006580.4 rs201367228 | F | 46-50 | 3 | N/A | Y | NL (radiopaque stones), Osteopenia, Ureter fissus<br>Normal plasma calcium (total 2.21; ionized 1.21)<br>Normal plasma magnesium (0.95)<br>Normal PTH (32)<br>Low 25-Vit D (16)<br>Normal 1,25-Vit D (84)<br>Hypercalciuria (6.7)<br>Normomagnesiuria (4.96) | Idiopathic NL | CLDN16-associated kidney stone disease |
| 5445 | *CLDN16* | AR | het | c.458A>G | p.Asn153Ser | LP | PS4, PM1, PP2, BS2, PP5 | 83/251.464/1 61/113.742/0 | NM_006580.4 rs201367228 | M | 36-40 | 2 | 90 COD, 10 COM | N | CaOx NL, Osteopenia<br>Normal plasma calcium (total 2.3; ionized 1.18)<br>Normal plasma magnesium (0.88)<br>Normal PTH (44)<br>Normal 25-Vit D (44)<br>Normal 1,25-Vit D (107)<br>Hypercalciuria (9.1)<br>Normal urine magnesium (5.3) | Idiopathic NL | CLDN16-associated kidney stone disease |

| | | | | | | | | | | | | | | | | |
|---|---|---|---|---|---|---|---|---|---|---|---|---|---|---|---|---|
| 5712 | *CLDN16* | AR | het | c.458A>G | p.Asn153Ser | LP | PS4, PM1, PP2, BS2, PP5 | 83/251.464/1 61/113.742/0 | NM_006580.4 rs201367228 | F | 21-25 | 3 | N/A | Y | NL, Osteopenia<br>Normal plasma calcium (total 2.34; ionized 1.18)<br>Normal plasma magnesium (0.79)<br>Normal PTH (32)<br>Normal 25-Vit D (52)<br>Normal 1,25-Vit D (127)<br>Hypercalciuria (7)<br>Normal urine magnesium (4) | Idiopathic NL | CLDN16-associated kidney stone disease |
| 5776 | *CLDN16* | AR | het | c.458A>G | p.Asn153Ser | LP | PS4, PM1, PP2, BS2, PP5 | 83/251.464/1 61/113.742/0 | NM_006580.4 rs201367228 | M | 26-30 | 2 | 100 COM | Y | CaOx NL, Osteopenia, Papillary calcification<br>Normal plasma calcium (total 2.36; ionized 1.17)<br>Normal plasma magnesium (0.84)<br>Normal PTH (53)<br>Normal 25-Vit D (67)<br>Normal 1,25-Vit D (111)<br>Hypercalciuria (5.7)<br>Normal urine magnesium (3.7) | Idiopathic NL | CLDN16-associated kidney stone disease |
| 5334 | *CLDN16*<br><br>**biallelic for CYP24A1* | AR | het | c.458A>G | p.Asn153Ser | LP | PS4, PM1, PP2, BS2, PP5 | 83/251.464/1 61/113.742/0 | NM_006580.4 rs201367228 | M | 21-25 | 3 | 1) 90 COM, 10 COD; 2) 80 COM, 20 COD | Y | CaOx NL, osteopenia, multiple kidney cysts.<br>Normal plasma calcium (total 2.48; ionized 1.27)<br>Normal plasma magnesium (0.72)<br>Low PTH (9)<br>Normal 25-Vit D (59)<br>Normal 1,25-Vit D (93)<br>Hypercalciuria (14.3) | Idiopathic NL, polycystic kidney disease | CLDN16-associated kidney stone disease |
| | **SLC4A1 – AD distal renal tubular acidosis (AD-dRTA)** | | | | | | | | | | | | | | | | |
| 5796 | *SLC4A1* | AR | het | c.1166G>A | p.Arg389His | LP | PP3s, PM2 | 3/251.344/0 1/113.670/0 | NM_000342.4 rs767059191 | M | 10-15 | 4 | 1) 90 COM, 20 COD<br>2) 100 COM<br>3) 90 COM, 10 COD<br>3) 90 COM, 10 COD | N | CaOx NL, DM II<br>Variable urine pH 5.7 – 7.0<br>Hypocitraturia (1.7)<br>Hyperoxaluria (564)<br>Normal plasma bicarbonate (24) and pH 7.35 | Idiopathic NL | AD-dRTA |

| | | | | | | | | | | | | | | | | | |
|---|---|---|---|---|---|---|---|---|---|---|---|---|---|---|---|---|---|
| | **SLC7A9 – Cystinuria, type B** | | | | | | | | | | | | | | | | |
| 5109 | SLC7A9 | AD/AR | 0/1 | C.544G>A | p.Ala182Thr | P | PM1s, PP2, PP5vs | 660/251.440/2 504/113.736/2 | NM_014270.5 rs79389353 | F | 21-25 | 4 | 80 COM, 20 COD | N | Recurrent CO KSD, Osteopenia lumbar spine, Intermittent Hypercalciuria (3.6-8.6), Hyperoxaluria (495-680 umol/d), Normal urine citrate (2.5-3.1), Urine cystine 4 mmol/mol Creatinine, Norm: <30) | Idiopathic NL | Cystinuria risk variant |
| 5323 | SLC7A9 | AD/AR | 0/1 | C.544G>A | p.Ala182Thr | P | PM1s, PP2, PP5vs | 660/251.440/2 504/113.736/2 | NM_014270.5 rs79389353 | M | 31-35 | 6 | N/A | N | Recurrent KSD, (no stone analysis), Hypersaluria (30g/d), low-low-normal urine Calcium (2.5-2.8), enteric Hyperoxaluria (2200-2360) post-Roux-en-Y Bypass, Low-Normal urine citrate (2.1-2.15) Urine cystine 5 mmol/mol Creatinine, Norm: <30) | Idiopathic NL | Cystinuria risk variant |
| 5409 | SLC7A9 | AD/AR | 0/1 | C.544G>A | p.Ala182Thr | P | PM1s, PP2, PP5vs | 660/251.440/2 504/113.736/2 | NM_014270.5 rs79389353 | F | 26-30 | 5 | 40 COD, 60 CHPD | N | Recurrent CA-P/CO KSD, prim. & sec. Hyperparathyroidism, Osteoporosis, Hypercalcemia (2.6-2.8, Ca-ion 1.33-1.44), Normal urinary calcium (3.7-4.8), Normal urine oxalate (299-403), normal urinary citrate (5.2) Urine cystine 9 mmol/mol Creatinine, Norm: <30) | Idiopathic NL | Cystinuria risk variant |
| 5211 | *SLC7A9* | AD/AR | het | c.313G>A | p.Gly105Arg | LP | PM1, PP2, PM5supp, PP3m, PP5 | 96/251.028/1 64/113.436/1 | NM_014270.5 rs121908480 | F | 41-45 | 4 | 90 COM, 10 COD | N | CaOx NL Hypercalciuria (5.3) Normal urine oxalate (237) Urine cystine (57 mmol/mol Creatinine) Norm: <30) | Idiopathic NL | Cystinuria risk variant |
| 5517 | *SLC7A9* | AD/AR | het | c.313G>A | p.Gly105Arg | LP | PM1, PP2, PM5supp, PP3m, PP5 | 96/251.028/1 64/113.436/1 | NM_014270.5 rs121908480 | M | 26-30 | 2 | 1) 70 COD, 30 CA 2) 10 COM, 60 COD, 30 CA; | Y | CaOx NL Hypercalciuria (8.6) Normal urine oxalate (297) Hypocitraturia (0.2) Urine cystine (59 mmol/mol Creatinine) Norm: <30) | Idiopathic NL | Cystinuria risk variant |

| | | | | | | | | | | | | | | | | |
|---|---|---|---|---|---|---|---|---|---|---|---|---|---|---|---|---|
| 5678 | *SLC7A9* | AD/AR | het | c.749+1G>C | p.? | LP | PVS1s, PM2, PP5 | 1/251434/0<br>1/113.738/0 | NM_014270.5<br>rs1060499787 | M | 41-45 | 3 | 80 Brushite, 10 COM, 10 CA | N | Brushite/CaOx NL<br>Hypercalciuria (7.5)<br>Normal urine oxalate (381)<br>Normal urine citrate (4.6)<br>Urine cystine (10 mmol/mol Creatinine) | Idiopathic NL | Cystinuria risk variant |
| | **SLC34A1 – Hypophosphatemic nephrolithiasis/osteoporosis-1 (NPHLOP1) / SLC34A1-related kidney stone disease** | | | | | | | | | | | | | | | | |
| 5143 | *SLC34A1* | AR | het | c.398C>T | p.Ala133Val | LP | PS3, PP3m, PP5 | 936/251.438/2<br>442/113.726/1 | NM_003052.5<br>rs148976897 | F | 31-35 | 3 | 70 COM, 15 COD, 15 CA | Y | CaOx NL, Osteopenia (lumbar spine & tibia),<br>Normal plasma calcium (total 2.23, ionized 1.21)<br>low plasma phosphate (0.79)<br>Normal PTH (27-42)<br>Normal 1,25-Vit D (101)<br>Hypercalciuria (6.5)<br>Hypocitraturia (1.6)<br>Hyperoxaluria (515-790)<br>Normal urine phosphate (13.3)<br>No glucosuria, normal urine cystine (trace), | recurrent NL | SLC34A1-related kidney stone disease |
| 5167 | *SLC34A1* | AR | het | c.398C>T | p.Ala133Val | LP | PS3, PP3m, PP5 | 936/251.438/2<br>442/113.726/1 | NM_003052.5<br>rs148976898 | M | 16-20 | 7 | 80 COM, 20 OCHPD | Y | CaOx NL, Normal BMD, normal plasma phosphate (1.04), low plasma phosphate at lowNa/Ca diet (0.73), increased FE-Phosphate (25%)<br>Normal plasma calcium (total 2.32-2.49, ionized 1.22-1.25)<br>Normal PTH (37-51)<br>Normal 25-Vit D (57)<br>Normal 1,25-Vit D (154-56)<br>Hypercalciuria (7.8-12.1)<br>Normal urinary citrate (3.25-4.5)<br>Hyperoxaluria (420-750)<br>Normal urine phosphate (25-39.4)<br>No glucosuria, normal urine cystine (trace), | recurrent NL | SLC34A1-related kidney stone disease |

| | | | | | | | | | | | | | | | | | |
|---|---|---|---|---|---|---|---|---|---|---|---|---|---|---|---|---|---|
| 5181 | *SLC34A1* | AD/AR | het | c.1130dup | p.Lys378GlnfsTer61 | LP | PVS1, PM2s | NF / NF | NM_003052.5 | F | 26-30 | 2 | 60 CA, 20 COD, 20 MAP | Y | CaP/CaOx NL, NC, CKD stage 3<br>Normal plasma calcium (total 2.46, ionized 1.22)<br>Normal plasma phosphate (1.05)<br>Normal PTH (23)<br>Normal 25-Vit D (64)<br>Normal 1,25-Vit D (61)<br>Hypocitraturia (0.2)<br>High urine oxalate (854)<br>Normal urine phosphate (25.4)<br>No glucosuria, normal urine cystine (4) | Idiopathic NL and NC, CKD | SLC34A1-related kidney stone disease |
| 5356 | *SLC34A1* | AR | het | c.398C>T | p.Ala133Val | LP | PS3, PP3m, PP5 | 936/251.438/2<br>442/113.726/1 | NM_003052.5<br>rs148976899 | M | 30-35 | 3 | 100 COM; 20 COM, 70 COD, 10 CA | Y | CaOx NL, Osteopenia, low plasma phosphate (0.52-0.62), Hypokalemia increased FE-Phosphate (20-30%)<br>Normal plasma calcium (total 2.41, ionized 1.18)<br>Normal PTH (33-45)<br>Low 25-Vit D (<12)<br>Normal 1,25-Vit D (89-100)<br>Normal urinary calcium (3.1-5)<br>Normal urinary citrate (2.3-3.9)<br>Normal urinary oxalate (290-380)<br>Normal urine phosphate (25-39.4)<br>No glucosuria, normal urine cystine (3), | recurrent NL | SLC34A1-related kidney stone disease |
| 5569 | *SLC34A1* | AR | het | c.398C>T | p.Ala133Val | LP | PS3, PP3m, PP5 | 936/251.438/2<br>442/113.726/1 | NM_003052.5<br>rs148976900 | M | 20-25 | 10 | 100 CO not diff | Y | CaOx NL, Normal BMD, low-normal plasma phosphate (0.77-1.07)<br>Normal plasma calcium (total 2.2.2.3, ionized 1.15-1.21)<br>Normal PTH (37-43)<br>Normal 25-Vit D (58-68)<br>Normal 1,25-Vit D (91-122)<br>Normal-high urinary calcium (2.8-6.3) | recurrent NL | SLC34A1-related kidney stone disease |

| | | | | | | | | | | | | | | | | | |
|---|---|---|---|---|---|---|---|---|---|---|---|---|---|---|---|---|---|
| | | | | | | | | | | | | | | | Low-normal urinary citrate (1.3-2.6)<br>Normal urinary oxalate (412-560)<br>Normal urine phosphate (15-30)<br>No glucosuria, normal urine cystine (3), | | |
| 5571 | *SLC34A1* | AR | het | c.398C>T | p.Ala133Val | LP | PS3, PP3m, PP5 | 936/251.438/2442/113.726/1 | NM_003052.5<br>rs148976901 | M | 60-65 | 2 | - | - | Recurrent struvite NL,<br>Normal plasma phosphate (1.06-1.16)<br>Normal plasma calcium (total 2.4, ionized 1.21)<br>Normal PTH (32)<br>Normal 25-Vit D (40-70)<br>Normal 1,25-Vit D (60-70)<br>Low urinary calcium (1.4-2.6)<br>Normal urinary citrate (2.3-3.6)<br>Normal urinary oxalate (308-470)<br>Normal urine phosphate (22-31)<br>No glucosuria, normal urine cystine (8), | recurrent NL | SLC34A1-related kidney stone disease |
| 5663 | *SLC34A1* | AD/AR | het | c.244G>T | p.Glu82Ter | P | PVS1, PM2, PP5 | 17/249.842/0<br>15/112.652/0 | NM_003052.5<br>rs377186926 | M | 36-40 | 1 | 1): 90 Brushite, 10 CA,<br>2): 100 Brushite | N | Brushite NL, NC, CKD stage 2<br>Normal plasma calcium (total 2.41, ionized 1.24)<br>Normal plasma phosphate (1.05)<br>Normal PTH (42)<br>Normal 25-Vit D (43)<br>Normal 1,25-Vit D (85)<br>Hypercalciuria (8.7)<br>Hypocitraturia (0.5)<br>Normal urine oxalate (180)<br>Normal urine phosphate (38.4)<br>No glucosuria, normal urine cystine (4), incomplete dRTA ($NH_4CL$ test) | Idiopathic NL and NC, CKD, incomplete dRTA | SLC34A1-related kidney stone disease |
| 5706 | *SLC34A1* | AR | het | c.398C>T | p.Ala133Val | LP | PS3, PP3m, PP5 | 936/251.438/2442/113.726/1 | NM_003052.5<br>rs148976902 | M | 10-15 | 1 | - | N | radiopaque NL, Normal BMD, | idiopathic NL | SLC34A1-related kidney stone disease |

| | | | | | | | | | | | | | | | | | |
|---|---|---|---|---|---|---|---|---|---|---|---|---|---|---|---|---|---|
| | | | | | | | | | | | | | | | normal plasma phosphate (1.11)<br>Normal plasma calcium (total 2.37-2.46, ionized 1.19-22)<br>Normal PTH (33-57)<br>Low 25-Vit D (<25-35)<br>Normal 1,25-Vit D (77-120)<br>Hypercalciuria (5.4-13)<br>Low-normal urinary citrate (1.3-4.2)<br>Normal urinary oxalate (205-306)<br>Normal urine phosphate (30-37)<br>No glucosuria, normal urine cystine (5), | | |
| 5719 | *SLC34A1* | AR | het | c.398C>T | p.Ala133Val | LP | PS3, PP3m, PP5 | 936/251.438/2442/113.726/1 | NM_003052.5 rs148976904 | F | 21-25 | 1 | - | Y | radiopaque NL, CKD Stage II, Normal BMD, normal plasma phosphate (0.93-1.15)<br>Normal plasma calcium (total 2.3-2.35, ionized 1.12-1.15)<br>Normal PTH (32)<br>Normal 25-Vit D (58)<br>Normal 1,25-Vit D (90)<br>Normal-high urinary calcium (3.6-7.2)<br>Low-normal urinary citrate (2.2-3.3)<br>Normal urinary oxalate (116-265)<br>Normal urine phosphate (28-32)<br>No glucosuria, normal urine cystine (3), | idiopathic NL | SLC34A1-related kidney stone disease |
| 5791 | *SLC34A1* | AR | het | c.398C>T | p.Ala133Val | LP | PS3, PP3m, PP5 | 936/251.438/2442/113.726/1 | NM_003052.5 rs148976905 | F | 31-35 | 1 | 60 CA, 40 CHPD | Y | CaP NL,<br>radiopaque NL, CKD Stage II, Normal BMD, Exclusion dRTA (NH4Cl & F/F-Test) Normal plasma phosphate (1.44)<br>Normal plasma calcium (total 2.18)<br>Hypercalciuria,<br>Low-normal urinary No glucosuria. | idiopathic NL, no dRTA | SLC34A1-related kidney stone disease |

| | | | | | | | | | | | | | | | | |
|---|---|---|---|---|---|---|---|---|---|---|---|---|---|---|---|---|
| | SLC9A3R1 | | het | c.458G>A | p.Arg153Gln | LP | PS3s, PM2, BP6supp | 462/251.340/1 381/113.668/8 | NM_004252.5 rs41282065 | | | | | | | | SLC9A3R1-related kidney stone disease |
| 5810 | *SLC34A1* | AR | het | c.1844del | p.Phe615SerfsTer67 | LP | PVS1s, PM2 | 2/242.762/0 2/111.942/0 | NM_003052.5 rs1318616234 | M | 31-35 | 3 | - | N | NL, Exclusion dRTA (NH4Cl & F/F-Test) Normal plasma phosphate (0.97), Normal plasma calcium (total 2.35, ionized 1.18) Normal PTH (42) Very low 25-Vit D (<25) Normal 1,25-Vit D (59) Low urinary citrate (No glucosuria. | Recurrent NL | SLC34A1-related kidney stone disease |
| 5860 | *SLC34A1* | AR | het | c.398C>T | p.Ala133Val | LP | PS3, PP3m, PP5 | 936/251.438/2 442/113.726/1 | NM_003052.5 rs148976906 | F | 21-25 | 1 | 70 COM, 30 COD | Y | CaOx NL, Exclusion dRTA Normal plasma calcium (total 2.2-2.27, ionized 1.13-1.17) Normal PTH (57) Low 25-Vit D (32-34) Normal 1,25-Vit D (83-120) Hypercalciuria (7.0-9.2) Normal urinary citrate (4.1-5.1) Normal-high urinary oxalate (214-508) Normal urine phosphate (28-32) No glucosuria, normal urine cystine (3), | idiopathic NL | SLC34A1-related kidney stone disease |
| 5917 | *SLC34A1* | AR | het | c.398C>T | p.Ala133Val | LP | PS3, PP3m, PP5 | 936/251.438/2 442/113.726/1 | NM_003052.5 rs148976907 | F | 41-45 | 1 | 70 COM, 30 COD | N | CaOx NL, low-normal plasma phosphate (0.70-0.82), Exclusion dRTA (NH4Cl-Test), Normal plasma calcium (total 2.2-2.3, ionized 1.23) Normal PTH (48) Low 25-Vit D (42) Normal 1,25-Vit D (128) Normal urinary calcium (2.5-5.2) | idiopathic NL | SLC34A1-related kidney stone disease |

| | | | | | | | | | | | | | | | | | |
|---|---|---|---|---|---|---|---|---|---|---|---|---|---|---|---|---|---|
| | | | | | | | | | | | | | | | Low-normal urinary citrate (1.97-3)<br>Normal urinary oxalate (300-330)<br>Normal urine phosphate (14-18)<br>No glucosuria. | | |
| 5925 | *SLC34A1* | AR | het | c.398C>T | p.Ala133Val | LP | PS3, PP3m, PP5 | 936/251.438/2 442/113.726/1 | NM_003052.5 rs148976908 | M | - | 3 | 20 COD, 40 CA, 40 OCHPD | Y | CaP/Ca-Ox NL, low-normal plasma phosphate (0.61-0.84),<br>Normal plasma calcium (total 2.35-2.4, ionized 1.25)<br>Normal PTH (37.5)<br>Low 25-Vit D (37)<br>Normal 1,25-Vit D (135)<br>Hypercalciuria (8-14)<br>Normal urinary citrate (2.8-4.6)<br>Normal-high urinary oxalate (380-720)<br>high urine phosphate (43-67)<br>No glucosuria. Low urinary cystine. | Recurrent NL | SLC34A1-related kidney stone disease |
| | **SLC34A3 – SLC34A3-related kidney stone disease / hypophosphatemic rickets with hypercalciuria (HHRH)** | | | | | | | | | | | | | | | | |
| 5021 | *SLC34A3* | AR | het | c.496G>A | p.Gly166Ser | LP | PP3s, PM2, PP5 | 10/249.840/0 6/112.568/0 | NM_001177316.2 rs200536604 | M | 31-35 | 3 | N/A | Y | NL (radiopaque stones)<br>Normal plasma calcium (total 2.35, ionized 1.23)<br>Low –normal plasma phosphate (0.77-0.95)<br>Normal PTH (46)<br>Normal 1,25-Vit D (94)<br>Hypercalciuria (8.9)<br>High urine phosphate (34.2) | idiopathic NL | SLC34A3-related kidney stone disease |
| 5045 | *SLC34A3* | AR | het | c.496G>A | p.Gly166Ser | LP | PP3s, PM2, PP5 | 10/249.840/0 6/112.568/0 | NM_001177316.2 rs200536604 | M | 26-30 | 3 | 100 CaOx | N | CaOx NL<br>Normal plasma calcium (total 2.48, ionized 1.23)<br>Normal plasma phosphate (1.13)<br>Normal PTH (48)<br>Normal 1,25-Vit D (103)<br>Hypercalciuria (15.5)<br>High urine phosphate (61) | idiopathic NL | SLC34A3-related kidney stone disease |
| 5048 | *SLC34A3* | AR | het | c.1055_1058dup | p.Val354ProfsTer240 | LP | PVS1, PM2 | NF / NF | NM_001177316.2<br>- | M | | 1 | N/A | Y | NL, NC<br>Normal plasma calcium (total 2.43, ionized 1.25) | NC with medullary sponge | SLC34A3-related kidney stone disease |

| | | | | | | | | | | | | | | | | | |
|---|---|---|---|---|---|---|---|---|---|---|---|---|---|---|---|---|---|
| | | | | | | | | | | | | | | | Low –plasma phosphate (0.74-1.01)<br>Normal PTH (29)<br>Normal 25-Vit D (71)<br>Normal 1,25-Vit D (71)<br>Normal plasma bicarbonate (28)<br>Hypercalciuria (7.1)<br>Normal urine phosphate (31)<br>Hypocitraturia (1.4)<br>High urine pH (7.5) | kidney, incomplete dRTA | |
| 5050 | *SLC34A3* | AR | het | c.1336-25_1336del | p.? | LP | PVS1s, PM2 | 1/244.272/0<br>1/107.636/0 | NM_001177316.2<br>rs775093177 | M | 51-55 | 4 | N/A | N | NL (radiopaque stones)<br>Normal plasma calcium (total 2.53, ionized 1.21)<br>Normal plasma phosphate (0.91-1.15)<br>Normal PTH (47)<br>Normal 1,25-Vit D (142)<br>Normal urine calcium (2.8)<br>Normal urine phosphate (20) | Idiopathic NL | SLC34A3-related kidney stone disease |
| 5070 | *SLC34A3* | AR | het | c.1242C>G | p.Tyr414Ter | LP | PVS1, PM2 | NF / NF | NM_001177316.2<br>rs949841477 | M | 21-25 | 1 | 80 COM 20 OCDP | Y | CaOx/CaP NL, osteopenia<br>Normal plasma calcium (total 2.54, ionized 1.26)<br>Normal plasma phosphate (0.95)<br>Normal PTH (18)<br>Normal 1,25-Vit D (144)<br>Hypercalciuria (6.3)<br>Normal urine phosphate (20) | Idiopathic NL | SLC34A3-related kidney stone disease |
| 5079 | *SLC34A3* | AR | het | c.1336-25_1336del | p.? | LP | PVS1s, PM2 | 1/244.272/0<br>1/107.636/0 | NM_001177316.2<br>rs775093177 | M | 41-45 | 4 | 1) 80 COD 20 COM<br>2) 80 UA, 10 COD 10 COM | Y | CaOx/UA NL, gout.<br>Normal plasma calcium (total 2.47, ionized 1.26)<br>Normal plasma phosphate (1.06-1.08)<br>Normal PTH (16)<br>Normal 1,25-Vit D (148)<br>Hypercalciuria (6.4)<br>High urine phosphate (38) | Idiopathic NL | SLC34A3-related kidney stone disease |
| 5102 | *SLC34A3* | AR | het | c.575C>T | p.Ser192Leu | P | PM2, PM5supp, PP3, PP5vs | 80/183.402/0<br>73/75.916/0 | NM_001177316.2<br>rs199690076 | M | 21-25 | 3 | 80 COD 10 COM 10 CA | Y | CaOx NL<br>Normal plasma calcium (total 2.58, ionized 1.29)<br>Normal plasma phosphate (0.81-0.95)<br>Normal PTH (37) | Idiopathic NL | SLC34A3-related kidney stone disease |

| | | | | | | | | | | | | | | | | |
|---|---|---|---|---|---|---|---|---|---|---|---|---|---|---|---|---|
| | | | | | | | | | | | | | | High-normal 1,25-Vit D (161)<br>Normal urinary calcium (3-4.3)<br>Normal urine phosphate (23-32)<br>Normal urine citrate (2.6-4.4) | | |
| 5164 | *SLC34A3* | AR | het | c.496G>A | p.Gly166Ser | LP | PP3s, PM2, PP5 | 10/249.840/0<br>6/112.568/0 | NM_001177316.2<br>rs200536604 | M | 15-20 | 1 | N/A | Y | NL, Normal plasma calcium (total 2.45, ionized 1.24)<br>Normal plasma phosphate (1.11-1.31)<br>Low PTH (13)<br>Normal 1,25-Vit D (132)<br>Hypercalciuria (9.4)<br>High urine phosphate (37) | Idiopathic NL | SLC34A3-related kidney stone disease |
| 5183 | *SLC34A3* | AR | het | c.496G>A | p.Gly166Ser | LP | PP3s, PM2, PP5 | 10/249.840/0<br>6/112.568/0 | NM_001177316.2<br>rs200536604 | M | 21-25 | 4 | 1) 35 COM, 30 COD, 35 CA<br>2) 10 COM, 85 COD, 5 CA | Y | CaOx/CaP NL<br>Normal plasma calcium (total 2.18)<br>Normal plasma phosphate (0.87)<br>Normal PTH (42)<br>Normal 1,25-Vit D (124)<br>Hypercalciuria (8.1)<br>Normal urine phosphate (24) | Idiopathic NL | SLC34A3-related kidney stone disease |
| 5312 | *SLC34A3* | AR | het | c.925+20_926-48del | p.? | LP | PP5vs, PM2, BP4 | 43/151532/0<br>23/67.800/0<br>(genomes) | NM_001177316.2<br>rs1554784044 | F | 6-10 | 4 | 100 CaOx | N | CaOx NL<br>Normal plasma calcium (total 2.42, ionized 1.21)<br>Low-normal plasma phosphate (0.72-0.91)<br>Normal PTH (22)<br>Normal 25-Vit D (68)<br>Normal 1,25-Vit D (141)<br>Hypercalciuria (6.2)<br>Normal urine phosphate (26) | Idiopathic NL | SLC34A3-related kidney stone disease |
| 5363 | *SLC34A3* | AR | het | c.1336-25_1336del | p.? | LP | PVS1s, PM2 | 1/244.272/0<br>1/107.636/0 | NM_001177316.2<br>rs775093177 | M | 26-30 | 2 | 60 COD 40 COM | Y | CaOx NL<br>Normal plasma calcium (total 2.33, ionized 1.22)<br>Low-normal plasma phosphate (0.75-0.89)<br>Normal PTH (31)<br>Low 25-Vit D (29)<br>Normal 1,25-Vit D (131)<br>Hypercalciuria (9.5)<br>Normal urine phosphate (30) | Idiopathic NL | stone disease |

| | | | | | | | | | | | | | | | | | |
|---|---|---|---|---|---|---|---|---|---|---|---|---|---|---|---|---|---|
| 5455 | *SLC34A3* | AR | het | c.496G>A | p.Gly166Ser | LP | PP3s, PM2, PP5 | 10/249.840/0 6/112.568/0 | NM_001177316.2 rs200536604 | M | N/A | 1 | N/A | N | NL (radiopaque stones) Normal plasma calcium (total 2.33, ionized 1.24) Normal plasma phosphate (0.84-1.24) Normal PTH (35) Low 25-Vit D (19) Normal 1,25-Vit D (58) Hypercalciuria (6.1) Normal urine phosphate (25) | Idiopathic NL | SLC34A3-related kidney stone disease |
| 5505 | *SLC34A3* | AR | het | c.304+2T>C | p.? | LP | PVS1s, PM2, PP5 | 9/250.156/0 8/112.704/0 | NM_001177316.2 rs201293634 | M | 16-20 | 3 | 80 COD 20 OCDP | N/A | CaOx/CaP NL Normal plasma calcium (total 2.23, ionized 1.17) Normal plasma phosphate (0.77-0-97) Normal PTH (24) Normal 25-Vit D (54) Normal 1,25-Vit D (84) Hypercalciuria (8.4) Normal urine phosphate (31) | Idiopathic NL | SLC34A3-related kidney stone disease |
| 5640 | *SLC34A3* | AR | het | c.1166dup | p.Gln390AlafsTer203 | LP | PVS1, PM2 | NF / NF | NM_001177316.2 | F | 46-50 | 2 | 1) 80 COM 20 COD 2) 60 COM 20 COD 20 CA | N | CaOx NL Normal plasma calcium (total 2.21, ionized 1.17) Normal plasma phosphate (0.82.1.04) Normal PTH (19) Low 25-Vit D (25) Normal 1,25-Vit D (63) Hypercalciuria (13.7) Normal urine phosphate (22) | Idiopathic NL | SLC34A3-related kidney stone disease |
| 5643 | *SLC34A3* | AR | het | c.496G>A | p.Gly166Ser | LP | PP3s, PM2, PP5 | 10/249.840/0 6/112.568/0 | NM_001177316.2 rs200536604 | M | 16-20 | 3 | 50 COM 30 COD 20 CA | Y | CaOx/CaP NL Normal plasma calcium (total 2.44, ionized 1.23) Low plasma phosphate (0.72-0.76) Normal PTH (18) Low 25-Vit D (37) Normal 1,25-Vit D (77) Hypercalciuria (13.5) High urine phosphate (45) | Idiopathic NL | SLC34A3-related kidney stone disease |
| 5828 | *SLC34A3* | AR | het | c.133del | p.Trp45GlyfsTer6 | LP | PVS1, PM2 | NF / NF | NM_001177316.2 - | M | 26-30 | 1 | 90 COD/10 COM | Y | CaOx NL Normal plasma calcium (total 2.39, ionized 1.19) Normal plasma phosphate (0.81-1.09) Normal PTH (45) | Idiopathic NL, dRTA excluded | SLC34A3-related kidney stone disease |

| | | | | | | | | | | | | | | | | | |
|---|---|---|---|---|---|---|---|---|---|---|---|---|---|---|---|---|---|
| | | | | | | | | | | | | | | | Low 25-Vit D (24)<br>Normal 1,25-Vit D (127)<br>Hypercalciuria (8.8)<br>High urine phosphate (33) | | |
| 5948<br>**NKSF** | *SLC34A3* | AR | het | c.496G>A | p.Gly166Ser | LP | PP3s,<br>PM2,<br>PP5 | 10/249.840/0<br>6/112.568/0 | NM_00117731<br>6.2<br>rs200536604 | F | N/A | 0 | N/A | N | No NL<br>Normal plasma calcium (total 2.4, ionized 1.22)<br>Normal plasma phosphate (0.93-1.02)<br>Normal PTH (39)<br>Normal 25-Vit D (67)<br>High 1,25-Vit D (361)<br>Hypercalciuria (8.1)<br>Normal urine phosphate (19) | **NKSF** | SLC34A3-related renal phosphate loss |
| 6026 | *SLC34A3* | AR | het | c.1623G>A | p.Trp541Ter | LP | PVS1s,<br>PM2,<br>PP5 | 3/157.820/0<br>3/63.926/0 | NM_00117731<br>6.2<br>rs762610288 | M | 56-60 | 5 | 1) 95 COM<br>5 COD2)<br>COM<br>3) COM | N | CaOx NL<br>Normal plasma calcium (total 2.51)<br>Normal plasma phosphate (0.85-1.01)<br>Normal PTH (54)<br>Normal 25-Vit D (70)<br>Normal 1,25-Vit D (88)<br>Hypercalciuria (5.3)<br>Normal urine phosphate (31) | Idiopathic NL | SLC34A3-related kidney stone disease |
| | **SLC9A3R1 – SLC9A3R1-related kidney stone disease / Hypophosphatemic nephrolithiasis/osteoporosis 2, (NPHLOP2)** | | | | | | | | | | | | | | | | |
| 5006 | SLC9A3R1 | | het | c.458G>A | p.Arg153Gln | LP | PS3s,<br>PM2,<br>BP6supp | 462/251.340/1<br>381/113.668/1 | NM_004252.5<br>rs41282065 | F | 21-25 | 4 | 20 COD, 60 CA, 20 MAP | N | CaP/CaOx NL, CKD G2-3, Medullary sponge kidneys with multiple calcifications, Normal BMD, Normal plasma calcium (total 2.3, ion: 1.18), Normal plasma phosphate (1.13-1.17)<br>Normal PTH (34-58)<br>Normal 25-Vit D (70)<br>High-normal 1,25-Vit D (160)<br>Hypercalciuria (5.3)<br>Normal urine phosphate (31) | idiopathic NL, medullary sponge kidney | SLC9A3R1-related kidney stone disease |
| 5259 | SLC9A3R1 | | het | c.458G>A | p.Arg153Gln | LP | PS3s,<br>PM2,<br>BP6supp | 462/251.340/1<br>381/113.668/3 | NM_004252.5<br>rs41282065 | M | 21-25 | 4 | 80 COM, 20 COD | Y | CaOx NL, Osteopenia lumbar spine, femur and tibia,<br>Normal plasma calcium (total 2.3-2.41, ion: 1.20) | idiopathic NL | SLC9A3R1-related kidney stone disease |

| | | | | | | | | | | | | | | | | |
|---|---|---|---|---|---|---|---|---|---|---|---|---|---|---|---|---|
| | | | | | | | | | | | | | | Normal plasma phosphate (0.95-1.19)<br>Normal PTH (49-61)<br>Normal 1,25-Vit D (85-111)<br>Hypercalciuria (4.9-7.5)<br>Normal urine phosphate (14-32), FE-P: 30%<br>Hyperoxaluria (470-1320)<br>Hypocitratiuria (1.0-2.5) | | |
| 5294 | SLC9A3R1 | het | c.458G>A | p.Arg153Gln | LP | PS3s, PM2, BP6supp | 462/251.340/1381/113.668/4 | NM_004252.5 rs41282065 | M | 51-55 | 3 | 80 COM, 20 COD; 70 Struvite, 30 CA | N | CaOx/CaP (Struvite,CA) NL , Osteopenia,<br>Normal plasma calcium (total 2.2-2.3, ion: 1.19)<br>Low-normal plasma phosphate (0.89-0.96)<br>Normal PTH (37)<br>Normal 25-Vit D (85)<br>Normal 1,25-Vit D (121)<br>Hypercalciuria (7-7.5)<br>High-normal urine phosphate (28-34), High-normale urinae oxalate (250-550)<br>Hypocitratiuria (0.2-2.3) | idiopathic NL | SLC9A3R1-related kidney stone disease |
| 5325 | SLC9A3R1 | het | c.458G>A | p.Arg153Gln | LP | PS3s, PM2, BP6supp | 462/251.340/1381/113.668/5 | NM_004252.5 rs41282065 | M | 26-30 | 4 | 80 COM, 20 COD; 80 COM, 20 COD | Y | CaOx NL Normal BMD<br>Normal plasma calcium (total 2.2, ion: 1.20), normal plasma phosphate (0.95-1.09)<br>Normal PTH (25-54)<br>Normal 25-Vit D (58-72)<br>Normal 1,25-Vit D (69-108)<br>Normal urinary calcium (1.7-2.6)<br>Normal urine phosphate (18-20), normal urine oxalate (300-350)<br>Low-normal urinary citrate (2.2-3.0) | idiopathic NL | SLC9A3R1-related kidney stone disease |
| 5500 **NKSF** | SLC9A3R1 | het | c.458G>A | p.Arg153Gln | LP | PS3s, PM2, BP6supp | 462/251.340/1381/113.668/6 | NM_004252.5 rs41282065 | M | N/A | 0 | N/A | N | No NL<br>Normal plasma calcium (total 2.25, ionized 1.18-1.22)<br>Normal plasma phosphate (1.0)<br>Normal PTH (29)<br>Normal 25-Vit D (95) | **NKSF** | SLC9A3R1-related renal phosphate loss |

| | | | | | | | | | | | | | | | | |
|---|---|---|---|---|---|---|---|---|---|---|---|---|---|---|---|---|
| | | | | | | | | | | | | | | Low-normal 1,25-Vit D (49)<br>Hypercalciuria (10.9)<br>High urine phosphate (51 | | |
| 5563 | SLC9A3R1 | het | c.458G>A | p.Arg153Gln | LP | PS3s, PM2, BP6supp | 462/251.340/1381/113.668/7 | NM_004252.5<br>rs41282065 | M | 31-35 | 10 | 100 COM; 80 COM 20 COD; 70 COM, 30 COD | N | CaOx NL, Normal BMD<br>Normal plasma calcium (total 2.2-2.3, ion: 1.18-1.23), normal plasma phosphate (0.93-1.05)<br>Normal PTH (30-35)<br>Low-Normal 25-Vit D (48)<br>Normal 1,25-Vit D (91-127)<br>Hypercalciuria (6.5-14.5)<br>High urine phosphate (25-57), Hyperoxaluria (700-800)<br>normal urinary citrate (2.3-4.7) | idiopathic NL | SLC9A3R1-related kidney stone disease |

**Normal/pathologic values laboratory parameters:**

Plasma calcium (mmol/l): 2.15-2.5
Plasma magnesium (mmol/l): 0.66-1.07
Plasma phosphate (mmol/l): 0.81-1.45
PTH (pg/ml): 15-65
25-Vit D (nmol/l): 50-135
1,25-Vit D (pmol/l): 48-190

Hypercalciuria (mmol/24h): >5
Hyperoxaluria (µmol/24h): >500
Hypocitraturia (mmol/24h): <1.65
Urine magnesium (mmol/24h): 3-5
Urine phosphate (mmol/24h): 13-32
